# Genome-wide meta-analysis of pneumonia suggests a role for mucin biology and provides novel drug repurposing opportunities

**DOI:** 10.1101/2021.01.24.21250424

**Authors:** William R. Reay, Michael P. Geaghan, 23andMe Research Team, Murray J. Cairns

## Abstract

Pneumonia remains one of the leading causes of death worldwide, particularly amongst the elderly and young children. We performed a genome-wide meta-analysis of lifetime pneumonia diagnosis (N=266,277), that encompassed the largest collection of cases published to date. Genome-wide significant associations with pneumonia were uncovered for the first time beyond the major histocompatibility complex region, with three novel loci, including a signal fine-mapped to a cluster of mucin genes. Moreover, we demonstrated evidence of a polygenic effect of common and low frequency pneumonia associated variation impacting several other mucin genes and *O*-glycosylation, further suggesting a role for these processes in pneumonia pathophysiology. The pneumonia GWAS was then leveraged to identify drug repurposing opportunities, including evidence that supports the use of lipid modifying agents in the prevention and treatment of the disorder. We also propose how polygenic risk could be utilised for precision drug repurposing through pneumonia risk scores constructed using variants mapped to pathways with known drug targets. In summary, we provide novel insights into the genetic architecture of pneumonia susceptibility, with future study warranted to functionally interrogate novel association signals and evaluate the suitability of the compounds prioritised by this study as repositioning candidates.

## INTRODUCTION

Pneumonia is characterised as an acute infection of the lung, with fluid filled alveoli and resultant restriction of oxygen intake being a key hallmark of its pathophysiology. There are a number of mechanisms known to cause pneumonia, however, bacterial or viral infection are the most common aetiologies^1^. Pharmacological intervention in pneumonia treatment is largely dependent on the infection source – for instance, bacterial induced pneumonia is treated with antibiotics. Yearly mortality rates worldwide from pneumonia remain high, even in the developed world where access to antibiotics and routine hospital care is usually unrestricted^2,3^. This necessitates a greater understanding of the mechanisms involved in pneumonia susceptibility and pathogenesis, which could be leveraged to identify novel treatments and inform the repositioning of existing drugs.

There has been considerable work undertaken to identify host factors which influence the onset and clinical course of pneumonia. Twin-based estimates of pneumonia heritability are still lacking, however, the heritability of death due to infections disease has been estimated as high as 40%, although further study is required ^4^. There have also been few studies which have used modern statistical genetics approaches to test for the existence of risk-increasing or protective alleles associated with pneumonia with sufficient power for surpassing genome-wide significance. Previously, a genome-wide association study of lifetime self-reported pneumonia diagnosis was published using participants obtained by 23andMe Inc. that identified a significant signal in the major histocompatibility complex (MHC) region on chromosome six^5^. We sought to increase statistical power to detect association signals by performing a genome-wide meta-analysis of self-reported pneumonia in the 23andMe cohort with SNP effects on a clinically ascertained pneumonia phenotype from the FinnGen consortium. The genetic architecture of pneumonia was further interrogated to identify novel risk genes and salient biological pathways, along with an estimate of genetic correlation with clinically significant phenotypes. These data were then considered in light of drug repurposing and provided support to a number of plausible repositioning opportunities.

## MATERIALS AND METHODS

### Genome-wide meta-analysis of pneumonia

The genome-wide meta-analysis was performed using two primary study cohorts from 23andMe Inc. and FinnGen (release 3), respectively, with full details of these cohorts and the meta-analysis procedure detailed in the supplementary methods. Summary statistics for a self-reported pneumonia phenotype were obtained from 23andMe as outlined by Tian *et al*.^5^. This self-reported phenotype was derived from an online survey of 23andMe customers about their medical history. In the final GWAS after quality control (QC), there were 40600 cases and 90039 controls. In addition, summary statistics for pneumonia were downloaded from the third release of the FinnGen database which combines genotype data from Finnish biobanks and digital health record data from Finnish health registries. The pneumonia phenotype chosen was *All pneumoniae* (J10 pneumonia), for which 15771 cases and 119867 controls were available for GWAS after QC.

The 23andMe and FinnGen summary statistics were meta-analysed using an inverse-variance weighted model with fixed effects as implemented by METAL version March 2011^6^. Firstly, we meta-analysed common variants, defined as sites with allele frequency > 1% in both the 23andMe and FinnGen cohorts. Variants were retained if they were available in both summary statistics and had an imputation quality that exceeded a minimum of 0.3 or a mean of 0.5 for variants not physically genotyped, resulting in 6888413 sites with an effect size estimate from the meta-analysis and a total sample size of 266277 individuals. Imputed rare variants available in both studies were subjected to a stricter filtering threshold for imputation quality such that only variants with a minimum imputation quality > 0.5 or a mean value > 0.7 were subjected to meta-analysis, with 834366 low frequency variants considered. In both instances, we further tested for heterogeneity between the contributing studies using Cochran’s *Q* test. Genome-wide summary statistics from the IVW meta-analysis were processed using the FUMA v1.3.6 (Functional Mapping and Annotation of Genome-Wide Association Studies) platform^7^. Genome-wide significant variants were characterised using the traditional *P* < 5 × 10^−8^ threshold, whilst suggestive significance was defined using a more lenient threshold of *P* < 1×10^−5^. We utilised the default settings for defining independent significant SNPs (r^2^ ≤ 0.6) and lead SNPs (r^2^ ≤ 0.1). The reference panel population for LD estimation was the UK biobank release 2b 10k White British panel, with LD blocks within 250 kb of each other merged into a single locus. We examined the effect of conditioning on two smoking GWAS via the multi-trait-based conditional & joint analysis (mtCOJO) framework implemented in GCTA v 1.93.2 beta (Supplementary Methods)^8,9^. For the *MUC5AC* lead SNP, we additionally performed a phenome-wide association study using the IEUGWAS database version 3.7.0 (https://gwas.mrcieu.ac.uk/), reporting SNPs using a conventional phenome-wide significance threshold of *P* < 1 × 10^−5^. Given the most significant association in this database was a GWAS of adult-onset asthma ^10^, we tested whether the association of SNPs proximal to *MUC5AC* was driven by the same underlying causal variant, assuming a single causal variant, via the *coloc* colocalisation methodology implemented in version 4 of the package ^11^. We also sought to replicate our results in two UK biobank (UKBB) pneumonia GWAS, specifically, a self-reported pneumonia phenotype performed in the automated GWAS pipeline by the MRC IEU group (ukb-b-4533, https://gwas.mrcieu.ac.uk/datasets/ukb-b-4533/), as well as a phecode ICD-10 UKBB GWAS performed in an automated series of GWAS by the authors of the SAIGE methodology (https://pheweb.org/UKB-SAIGE/pheno/480) ^12^.

### Estimation of SNP-based heritability

SNP based heritability was computed using LD score regression (LDSR) with 1000 genomes phase 3 LD scores and weights^13^. We converted the heritability estimate to the liability scale assuming the population prevalence of pneumonia as that of pneumonia in the FinnGen dataset (12.61%), as well as a more conservative estimate based on ICD-10 diagnosed pneumonia in the UK biobank (UKBB) sample (3.20% - Supplementary Methods).

### Finemapping genome-wide significant loci

We finemapped the three-novel genome-wide significant loci outside of the MHC region by using a method which leverages asymptotic Bayes’ factors (ABF) to estimate credible sets under the assumption of a single causal variant^14^. Specifically, we utilised Wakefield’s method to approximate ABFs assuming a prior variance of 0.2^2^, which reflects the belief that the confidence intervals of estimated variant effect sizes expressed as odds ratios ranging from around 0.68 to 1.48. Given that the posterior probability for causality of each variant is proportional to its Bayes’ factor, these can be summed until a prespecified probability (ρ) is reached, thus, constituting a ρ set of putative causal variants. In this study, we derived 95% credible sets. A single causal variant was assumed such that we did not have to account for LD between variants, which has been demonstrated to be problematic in finemapping studies which prespecify more than one causal variant using references external to the GWAS like the 1000 genomes project panel^15^.

### Gene-based and gene-set association

Common variant (MAF > 0.01) SNP-wise *P* values were aggregated at gene-level using MAGMA v1.07b^16^, as described in the supplementary methods. The Bonferroni threshold for genic association was *P* < 2.68 × 10^−6^, accounting for the number of genes tested. Moreover, gene-based *P* values were leveraged for gene-set association using 1379 hallmark and canonical gene-sets from the Molecular Signatures Database (MSigDB)^17^. Rare variants (MAF < 0.01) were also aggregated at gene-level by leveraging the properties of the Cauchy distribution (Supplementary Methods). Code for the Cauchy combination test was obtained from (https://github.com/yaowuliu/ACAT) and outlined by Liu and Xie^18,19^. The MAGMA approach for common variants accounts for dependency between *P* values by estimating their covariance as a function of pairwise LD in a population sample – however, there are methodological challenges with this approach for rare variants and likely much larger samples would be required for accurate estimation of dependency between rare variants, if any exists^20,21^. Therefore, we employed Cauchy transformation to combine *P* values as it guards against type I error inflation due to potential unknown covariance between rare variants (Supplementary Methods). In addition, we constructed a model for rare variant gene-set association analogous to the MAGMA approach for common variants that leverages gene-based *Z* values (probit transformation of *P*). The same collection of pathways from MSigDB were considered, with genic *Z* values regressed against a binary indicator of set membership (*β*_S_), covaried for logarithmically transformed gene-length, and rare variant count per gene. A one-sided test was performed for *β*_S_, such that the null hypothesis is *β*_S_ = 0 and the alternative *β*_S_ > 0. Only gene-sets with rare variants overlapping at least 5 genes were retained.

### Transcriptome-wide association studies of pneumonia

A transcriptome-wide association study (TWAS) of pneumonia was performed using the FUSION package^22^. We utilised GTEx v7 SNP weights from three tissues that would be plausibly involved in the pathophysiology of pneumonia (whole blood, lung, and spleen). We corrected for the number of *cis*-heritable genes outside the MHC region for which a TWAS *Z* could be calculated in each tissue (Supplementary Methods). For transcriptome-wide significant genes, we tested whether the expression and pneumonia-associated signal displayed statistical colocalisation as encompassed by the SNP weights with the *coloc* package as implemented by FUSION^11^. In addition, we probabilistically finemapped transcriptome-wide significant regions using the FOCUS approach to derive a credible set of putative causal genes, as described previously^23^. We utilised the default Bernoulli prior (*p* = 1 × 10^−3^) and chi-square prior variance (*nσ*^2^ = 40) to approximate Bayes’ factors for each gene, and thus, derive the posterior inclusion probabilities (*PIP*) for each gene to be causal given its observed TWAS *Z*.

### Genetic correlation and causal inference

We estimated genetic correlation between pneumonia and 180 high quality, European ancestry GWAS using LDSR as implemented by the LDhub application^24^. For Bonferroni significant genetic correlation estimates, we constructed a latent causal variable (LCV) model using the most significant trait from each LDhub phenotypic category and pneumonia to evaluate evidence for genetic causality between traits, as outlined extensively elsewhere^25–27^. A strong estimate of the posterior genetic causality proportion (GCP) was defined as significantly different from zero (one sided *t*-test) and an absolute GCP estimate > 0.6. Weak GCP estimates close to zero for genetically correlated traits imply that their relationship is potentially mediated by horizontal pleiotropy, whereby there are shared pathways, but the two traits do not likely exhibit vertical pleiotropy by acting within the same pathway. We additionally evaluated evidence for a causal relationship between HDL cholesterol and pneumonia by constructing a multivariable Mendelian randomisation (MR) model using the TwoSampleMR package^28^. This multivariable model leveraged genetic instrumental variables (IV) from three highly biologically interconnected lipid traits (LDL, HDL, and triglycerides) and estimated the effects of these IVs on the outcome conditioned on their association with the other two lipid classes^29^.

### Genetically informed drug repurposing

We implemented three strategies to propose drug repurposing candidates: i) single loci drug-gene matching, ii) genetic correlation and/or evidence of a putative causal relationship between a biochemical trait that could be targeted by an approved drug, and iii) precision drug repurposing using the polygenic scoring orientated *pharmagenic enrichment score* (PES) approach. Full details of these analyses are described in the supplementary methods. We utilised a panel of 50 biochemical GWAS performed by the Neale lab from the UK biobank which had high or medium confidence estimates of SNP heritability that were significantly different from zero (http://www.nealelab.is/uk-biobank) and estimated genetic correlation and the posterior mean GCP for trait pairs that survived Bonferroni correction. We sought to replicate the results of the multivariable MR for the three lipid classes (LDL, HDL, and Triglycerides) using the UK biobank GWAS. MR was then performed to evaluate further evidence for a causal effect between gamma-glutamyltransferase (GGT) and pneumonia, as well as triglycerides and pneumonia (univariable estimate). Specifically, we defined independent, non-palindromic genome-wide significant variants as IVs and constructed four MR models with differing underlying assumptions (two inverse-variance weighted estimators with fixed or multiplicative random effects, weighted median estimator, weighted mode estimator, and MR-Egger)^30–33^. A series of sensitivity analyses to evaluate statistical evidence for confounding pleiotropy was then undertaken as outlined in the supplementary methods^32– 35^.

The PES framework is based on the postulation that an enrichment of genetic risk within a biological pathway with known drug targets may be an impetus to repurpose a drug which modulates that pathway for individuals who carry the high genetic load mapped to the pathway, as described elsewhere^36,37^. Specifically, we identify druggable pathways with an enrichment of common variant associations relative to the rest of the genes tested and construct pathway-based risk scores for these gene-sets (Supplementary Methods). We utilised the UK biobank (UKBB) cohort to test the association between a pneumonia PES and pneumonia phenotypes recorded for these participants^38,39^. These analyses are described in detail in the supplementary methods. Briefly, we retained 336,896 unrelated white British ancestry participants and 13,568,914 autosomal variants that survived a series quality control steps, including, imputation quality filtering (INFO > 0.8), MAF > 1 × 10^−4^, call rate > 0.98, and filtering strong deviations from the Hardy-Weinberg equilibrium. Self-reported pneumonia diagnosis and ICD-10 codes from hospital inpatient records were used to construct the pneumonia phenotype (Supplementary Methods). There were 10,540 individuals from the genotyped subset of the cohort included in the PES calculation with a primary or secondary diagnosis using the ICD-10 primary or secondary diagnosis codes relevant to pneumonia. In the strict phenotype definition, we defined cases as those satisfying ICD-10 criteria, and controls as all those who did not have one of those codes recorded along with any individual who self-reported pneumonia without a pneumonia ICD-10 code (N_Controls_ = 320,213). Individuals who self-reported pneumonia but were not assigned a relevant ICD-10 code were excluded from the study cohort in this strict configuration. In the broad-phenotype definition, pneumonia cases were individuals with a relevant ICD-10 code or a self-reported lifetime pneumonia diagnosis (N = 15,138). In other words, the strict definition only included individuals with a pneumonia ICD-10 code. The PES was then constructed using common, autosomal variants outside of the MHC region mapped to genes in that pathway with PRsice2 assuming an additive model, with further details provided in the supplementary methods^40^. The *P* value threshold of including variants in the PES was the same as what was used to identify the gene-set. A genome wide PRS for pneumonia susceptibility was also constructed in an analogous fashion.

We explored the phenotypic relevance of pneumonia PES profiles using a random subset of the UKBB (N ∼ 10,000) for which a multiplex assay was performed to quantity immunoglobulin G (IgG) antibody response to a series of antigens for infectious agents selected for study, as outlined elsewhere^41^. The two phenotypes of interest here were a binary indicator of *seropositivity* for 14 infections with seroprevalence > 5% in our genotyped subset of the cohort, and amongst seropositive individuals, a continuous measure of antibody response (mean fluorescence intensity) to each antigen for that infection – termed *seroreactivity*. The correlation between PES or PRS and seroreactivity was assessed by linear regression, whilst logistic regression was utilised for seropositivity. Both models were covaried for sex, age, age^2^, ten SNP derived principal components, genotyping batch, and two QC metrics related to the antibody assay (Supplementary Methods). A heatmap of the regression *t* statistics was constructed using the ComplexHeatmap package^42^

## RESULTS

### Novel common and rare variant loci associated with pneumonia

We performed a genome-wide meta-analysis of pneumonia using common and rare (MAF < 0.01) overlapping variants from 23andMe and FinnGen release three, with 6,888,413 and 834,366 common and low frequency sites tested, respectively. We estimated the SNP based heritability as approximately 3.24% on the liability scale (Fig. 1b), using the incidence of pneumonia in the FinnGen cohorts as the population prevalence (12.67%), although we acknowledge the population prevalence of pneumonia is difficult to quantify. As a result, we re-estimated *h*^2^ using a more conservative population prevalence value based on phenotype data from the UK biobank (3.20%), resulting in a lower estimate of *h*^2^_SNP_ = 0.0213. The point estimate of SNP-based *h*^2^ was higher in the 23andMe cohort. *h*^2^_SNP_ = 0.054, although the estimate was more precise in the meta-analysis than in 23andMe and FinnGen alone: *Z*_Meta_ = 9.26, *Z*_23andMe_ = 7.44, and *Z*_FinnGen_ = 4.12. In line with previous comparisons between self-reported and clinically ascertained phenotypes, the heritability estimate was lower in FinnGen than 23andMe. There was some evidence of test statistic inflation when visualised as a *QQ* plot (Supplementary Figure 1), however, the proportion of the polygenic signal in the meta-analysis attributed to model misspecification and/or confounding was around 11% (LDSR ratio = 0.1145), whilst the mean *χ*^2^ was large enough to estimate heritability (1.11).

**Figure 1.**
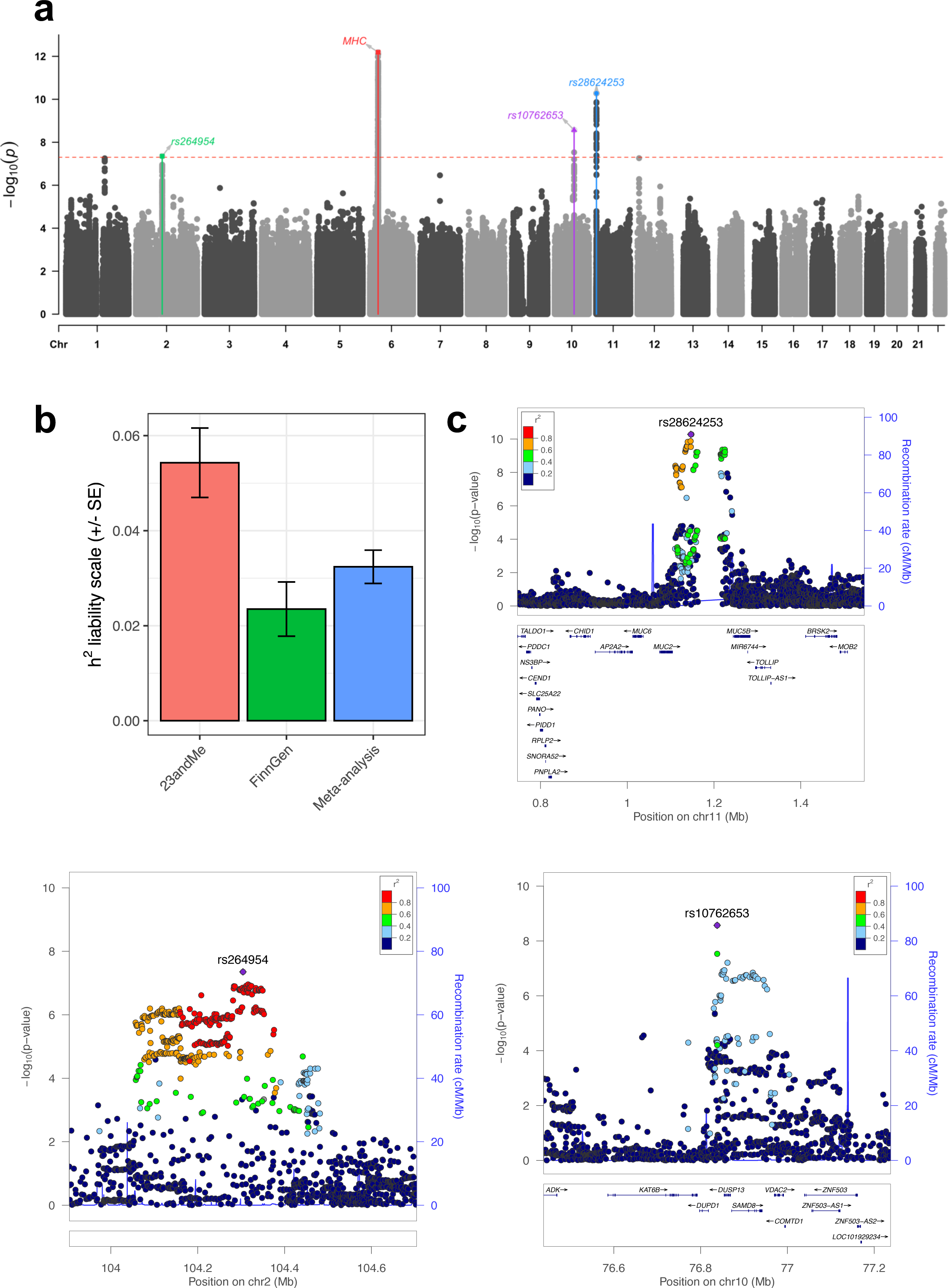
Genome-wide meta-analysis of pneumonia susceptibility. (**a**) Manhattan plot of common variant GWAS for pneumonia, as is usual practice, each point is the - log_10_ *P* value of a variant for association with pneumonia, with the red dotted line indicative of genome-wide significance (*P* < 5 × 10^−8^). Lead SNPs are highlighted and labelled on the plot, except for the MHC locus which we denote as “MHC” due to its complexity. (**b**) Estimates of SNP-based heritability (*h*^2^) on the liability scale for the 23andMe and FinnGen cohorts individually, as well as the using the inverse-variance weighted effects meta-analysis of the two cohorts. The error bars represent the standard error of *h*^2^. (**c**) Region plots for the three-novel genome-wide significant loci outside of the MHC region, the LD for each variant with the lead SNP estimates from the 1000 genomes phase III European reference set was utilised to colour the points.

There were four common genomic loci that surpassed the conventional genome-wide significance threshold (*P* < 5 × 10^−8^, Table 1, Fig. 1a,c, Supplementary Fig. 2-5). The effect sizes of these common variant signals were small in accordance with expectation, with each SNP increasing or decreasing the odds of pneumonia by around 5%. Unsurprisingly, the most significant signal spanned the major histocompatibility complex (MHC) region, which has been previously published as associated with pneumonia in the 23andMe cohort^5^. Due to the complexity of this region, we define the MHC signal as a single locus, with the minor allele of the lead common SNP associated with a small reduction in the odds of pneumonia (OR = 0.94 [95% CI: 0.92, 0.96], *P* = 6.44 × 10^−13^). The most significant novel common signal in this study was a region located on chromosome 11, with the lead SNP (rs28624253) located upstream of the *MUC5AC* gene that encodes a mucin protein, with three other genes that encode a mucin protein within 400 kilobases of the lead SNP *MUC6, MUC2*, and *MUC5B*. Importantly, rs28624253 was similarly associated in the 23andMe (*P* = 2.65 × 10^−6^) and FinnGen (*P* = 3.42 × 10^−6^) cohorts and there was no appreciable evidence for differences in population structure between the input GWAS driving this signal. Mucins are heavily glycosylated proteins that play a number of important roles, particularly in relation to the maintenance of mucosal barriers^43^. Mucin genes are known the exhibit somewhat pervasive genomic complexity and evidence of heterogeneity between populations, to ensure that this signal is not just an artefact of this, we performed a phenome-wide association study of the lead SNP and found that this variant was associated with only relevant phenotypes to pneumonia. Specifically, it was linked to adult-onset asthma, self-reported regular cough and mucus, and eosinophil count at a conventional phenome-wide significance threshold (*P* < 1 × 10^−5^). Given rs28624253 was associated with adult-onset asthma at a more stringent level of genome-wide significance, we tested whether the mucin signal observed for pneumonia and adult-onset asthma were driven by the same underlying causal variant and found strong evidence to support this hypothesis (posterior probability > 90%). We caution that this assumes the existence of a single causal variant, which may be unrealistic given the complexity of this region. As we visualise in supplementary figure 6, if one utilises a more conservative prior probability of a shared causal variant than there is some evidence that there is a different underlying causal variant but that the locus is still associated with both traits. It should also be noted that the odds increasing allele for pneumonia (*G*) is perhaps counterintuitively associated with decreased risk of asthma, which suggests a multifaceted biological mechanism which may be related to the role of mucins in the airway.

**Table one:** Lead SNPs within common genome-wide significant loci associated with pneumonia.

| Lead SNP | Locus | EA/NEA | EAF (NFE) | EAF (FIN) | OR | 95% CI | $P_{\text{GWAS}}$ | $P_{\text{Het}}$ |
| --- | --- | --- | --- | --- | --- | --- | --- | --- |
| rs9268966 | MHC | G/A | 0.26 | 0.33 | 0.94 | 0.92, 0.96 | $6.44\text{e}^{-13}$ | 0.04 |
| rs28624253 | chr11:1110395-1232702 | G/A | 0.37 | 0.41 | 1.05 | 1.04, 1.07 | $5.27\text{e}^{-11}$ | 0.44 |
| rs10762653 | chr10:76815686-76993015 | G/A | 0.16 | 0.18 | 1.06 | 1.04, 1.08 | $2.70\text{e}^{-9}$ | 0.61 |
| rs264954 | chr2:104056454-104380545 | T/C | 0.46 | 0.45 | 0.96 | 0.95, 0.98 | $4.44\text{e}^{-8}$ | 0.53 |

Common ( $\text{MAF} > 0.01$ ) lead SNPs were defined as independent SNPs ( $r^2 < 0.1$ ) within each genomic locus. The effect allele (EA) and non-effect allele (NEA) was reported for this table such that the effect allele was the minor allele. The effect allele frequency (EAF) is denoted in gnomAD v2.1.1 for non-Finish Europeans (NFE) and Finns (FIN). All odds ratio and their respective confidence intervals were calculated relative to the EA. Heterogeneity of effect between the 23andMe and FinnGen cohorts was tested using Cochran’s $Q$ , with the $P$ value of that test reported here ( $P_{\text{Het}}$ ). Due to the complexity of locus 1 (MHC), we report only a single common SNP for this locus. Locus coordinates in in hg19 assembly.

The third most significant locus encompassed several genes including *SAMD8, DUSP13*, and *VDAC2*, whilst locus four was largely intergenic, with some variants overlapping long-noncoding RNAs with limited annotation information (Supplementary Text). There were also two loci that almost surpassed genome-wide significance; specifically, a locus physically mapped to the interleukin-6 receptor region (*IL6R* – lead SNP: rs12730036, *P* = 5.61 × 10^−8^), and a region on chromosome 12 where the lead SNP is mapped to an intron of *TNFRSF1A*, which encodes a tumour necrosis factor receptor (lead SNP: rs1800693, *P* = 5.5 × 10^−8^). We integrated functional genomic and annotation data to prioritise candidate genes from the novel genome-wide significant loci in this study, as described in the supplementary text. We found that genes in the non-MHC loci implicated by at least two lines of evidence were enriched in phenotypically relevant pathways such as mucus layer, lung fibrosis, and *O*-linked glycosylation of mucins (Supplementary Figure 5, Supplementary Text).. Interestingly, despite the meta-analysis combining a self-reported phenotype with clinically ascertained pneumonia diagnoses, only the lead SNP in the MHC locus demonstrated any significant heterogeneity in the effect sizes between the two cohorts (Cochran’s *Q, P*_Het_ = 0.04, Table 1, Supplementary Text, Supplementary Figure 7).

The novel genome-wide significant loci outside the MHC region were finemapped to estimate a 95% credible set of plausible causal variants assuming a single causal variant in each region (Supplementary Tables 1-3). The variants encompassed by the 95% credible set for the rs28624253 locus were all proximally upstream/downstream of *MUC5AC*, or within the gene itself, supporting the relevance of this mucin gene for that association signal. The second most significant novel pneumonia associated locus discovered in this study (rs10762653 lead SNP) had a smaller credible set, with the lead SNP having a considerably high posterior probability than the remaining credible set SNPs (*PP* = 0.676). Interestingly, the lead SNP for this locus has low estimated LD with other proximal genome-wide significant SNPs (Fig 1c), suggesting the existence of several causal variants that cannot be accounted for by this method. Finally, the intergenic region spanned by the third novel genome-wide significant locus yielded a large credible set of over 200 variants and, as a result, further functional interrogation is required to mechanistically interpret this locus.

Smoking status was not included as a covariate in the respective GWAS meta-analysed, and thus, we sought to investigate whether genetic variants associated with smoking may confound our findings in this GWAS. Specifically, we genetically conditioned common variant associations on their effect size from a GWAS of smoking initiation and smoking heaviness using mtCOJO^8^. The effect sizes of the lead SNPs from the novel genome-wide significant loci were not greatly attenuated after conditioning on either of the smoking phenotypes and remained genome wide significant, with the exception of the locus tagged by rs264954 (Conditioned on smoking initiation: *P* = 7.15 × 10^−7^; Conditioned on smoking heaviness: *P* = 1.05 × 10^−7^). Furthermore, there was a slight reduction in the SNP heritability estimate on the liability scale, although this only amounted to a less than 0.5% difference after conditioning on either smoking phenotype – *h*^2^_Conditioned on smoking initiation_ = 3.08%, *h*^2^_Conditioned on smoking heaviness_ = 2.95%.

We also uncovered a genome-wide significant association between a rare intergenic variant in the MHC region and pneumonia - rs11962863, OR = 1.59 [95% CI: 1.44, 1.74], *P* = 1.15 × 10^−9^. This relatively large effect allele, however, did display statistically significant heterogeneity in its effect between the two cohorts (*P* = 8.20 × 10^−5^). This locus is considerably rarer in the Finnish population (AF = 5.8 × 10^−4^) than non-Finnish Europeans in gnomAD (AF = 2.9 × 10^−3^), which may account for its larger effect size in the FinnGen cohort. Due to the complexity of recombination and linkage in the MHC locus, the functional consequences of this variant remains difficult to interpret at an individual level without considering the local genomic context of affected individuals, such as HLA type. We also detected six additional regions with rare variants that surpassed suggestive significance for association with pneumonia (*P* < 1 × 10^−5^, Supplementary Table 4).

We sought to replicate our genome-wide significant and suggestively associated common loci using two GWAS from the independent UK Biobank cohort – specifically, we utilised two automated GWAS that encompassed a self-reported pneumonia phenotype (N_Case_ = 6572, N_Controls_ = 456,361) and ICD-10 derived pneumonia diagnoses (N_Case_ = 10,059, N_Controls_ = 398,538). We investigated both phenotyping approaches given our GWAS was a meta-analysis of self-reported and clinically ascertained data. In the self-reported pneumonia UKBB GWAS, we found that no SNPs replicated at genome-wide significance, however, the *MUC5AC* lead SNP was nominally associated in the same direction (*β* = 0.001, *SE* = 2.6 × 10^−3^, *P* = 0.022), with the MHC lead SNP also was also nominally significant (*P* = 0.03) and the remaining two lead SNPs demonstrated no significant evidence of replication. The ICD-10 phenotype GWAS in the UKBB did not replicate any of our non-MHC genome-wide significant SNPs at even nominal significance, although MHC SNPs were found to be nominally significant. It should be noted that a limitation of both GWAS is that they focused only on either the self-reported or clinically ascertained phenotype in the UKBB, meaning some controls plausibly would have had pneumonia, and thus, decreasing power. Moreover, the effective sample sizes (N_eff_) of these UKBB GWAS were markedly smaller than ours (177749 in the current discovery meta-analysis versus 25915 and 39246, respectively).

We also considered two very recent smaller sample-size pneumonia GWAS without publicly available summary statistics to see if we could replicate their findings. Firstly, Chen *et al*. performed a GWAS of pneumonia susceptibility and severity in the Vanderbilt University Biobank (BioVU, N_Case_ = 8889, N_Controls_ = 60,767, N_eff_ = 31019), European ancestry cohort)^44^. They found that a genome-wide significant common signal in Europeans associated with pneumonia severity, with the lead SNP rs10786398 nominally associated in our meta-analysis: *β* = -0.029, *SE* = 0.001, *P* = 2.5 × 10^−4^, whilst we were unable to replicate the significant rare-variant association signal from that study as the variant was not available in our analyses. Moreover, a meta-analysis of a smaller previous FinnGen release and ICD-10 (N_eff_ = 94584) derived pneumonia in the UKBB found two genome-wide significant index SNPs in the 15q15.1 region that were directionally consistent in our analyses, although not statistically significant, with a trend observed for rs76474922: *β* = 0.025, *SE* = 0.01, *P* = 0.08 ^45^. The SNP-based heritability estimate from that study also closely mirrored ours (3.3% on the liability scale), supporting the reliability of this study’s estimate in a larger sample.

### Gene and gene-set association further supports a role for mucin biology in pneumonia

We performed gene and gene-set association to investigate associations with pneumonia beyond univariable SNP-phenotype relationships (Supplementary Tables 5,6). Six genes outside of the MHC region were significant in the meta-analysis after the application of multiple-testing correction (*MUC5AC, MUC5B, DUPD1, SAMD8, DUSP13*, and *TOX*), all of which were within genome-wide significant loci except for the *TOX* gene (*P* = 1.26 × 10^−6^), which plays a role in T cell persistence during response to a pathogen^46,47^. Gene-set association revealed a single gene-set for which its member genes were enriched with pneumonia associated common variation relative to all other genes tested: *Termination of O-glycan biosynthesis* – *β* = 0.89, *SE* = 0.21, *P* = 1.05 × 10^−5^, *q* = 0.01. The strongest gene-based signal in this pathway was accounted for by two mucin genes that span part of the genome-wide significant loci on chromosome 11 (*MUC5AC, MUC2*), whilst there were four other mucin genes within this set that displayed a nominal gene-based association (*P* < 0.05) outside that region (*MUC15, MUC16, MUC12*, and *MUC17*). The signal from this gene-set remained relatively robust upon using a more conservative definition of the genic boundaries for SNP to gene annotation during gene-based association: *β* = 0.66, *SE* = 0.19, *P* = 3.04 × 10^−4^. Rare variants were then subjected to gene-based association by leveraging the properties of the Cauchy distribution, such that covariance between *P* values did not need to be estimated. No genes surpassed Bonferroni correction, considering genes with at least two rare variants. The most significant gene found upon testing all genic rare variants was *ZNF19, P* = 6.3 × 10^−5^, whilst *FREM1* was the top gene (*P* = 8.16 × 10^−4^) considering variants annotated as exonic only (Supplementary Tables 7,8). Furthermore, we utilised the gene-based results to construct a competitive rare variant gene-set association model and tested the same gene-sets as in the common variant analyses (Supplementary Tables 9,10). While there were no gene-sets that survived multiple-testing correction utilising all genic rare variants or those with only exonic annotated sites, we found some support for B lymphocyte antigen response in the rare variant architecture of pneumonia, as the most significant association from the rare variant model was with the *B cell antigen receptor* pathway - *β* = 0.59, *SE* = 0.19, *P* = 7.7 × 10^−4^. Interestingly, there was also a nominal rare variant signal amongst pathways related to carbohydrate metabolism (*Metabolism of carbohydrates* – *P* = 6.22 × 10^−3^, *Glycosaminoglycan degradation* – *P* = 0.02) and glycosylation (*N*-*glycan biosynthesis* – *P* = 0.01), although these results must be interpreted cautiously given, they do not survive multiple testing correction.

A transcriptome-wide association study was then undertaken to identify genes for which genetically predicted expression was correlated with pneumonia susceptibility. We selected SNP weights from three tissues which are plausibly biologically relevant to pneumonia pathophysiology: lung, whole blood, and spleen. After applying Bonferroni correction to the number of tests within each tissue outside the MHC individually, significant correlation was observed between decreased predicted expression of *VDAC2* and pneumonia (*Z* = -4.39, *P* = 1.15 × 10^−5^, Supplementary Table 11) using spleen tissue as the prediction model. The association between *VDAC2* and pneumonia was also supported by SNP weights from the lung and whole blood, although these did not surpass the Bonferroni threshold: *P*_Lung_ = 5.52 × 10^−4^, *P*_Whole blood_ = 9.97 × 10^−4^. We tested a related, but distinct hypothesis by assessing evidence for colocalisation between the GWAS signal spanning *VDAC2* and the SNP weights from the three tissues utilised to construct the model of genetically predicted *VDAC2* expression. Colocalisation assumes a biologically conservative parameter of a single shared causal variant that underlies the relationship between *VDAC2* expression and pneumonia. We observed heterogeneity between the three tissues, with moderately strong evidence of colocalisation between *VDAC2* expression and SNP weights from whole blood (*PP*_H4_ = 0.848), however, using lung and spleen expression SNP weights there was no strong evidence (*PP* > 0.8) for any of the five colocalisation hypotheses. In lung, there was evidence for an association between *VDAC2* lung expression SNP weights and pneumonia (*PP*_H3_ = 0.568, *PP*_H4_ = 0.363), although we were not able to clearly determine whether there were two independent SNPs driving the association (H3) or a shared variant (H4). Interestingly, there was moderate support in the model leveraging the spleen SNP expression weights that this signal was only associated with pneumonia and not *VDAC2* expression (*PP*_H2_ = 0.643), which could be driven by the spleen model of genetically regulated expression being somewhat less predictive (*R*^2^ = 0.038, best linear unbiased prediction), than in blood (*R*^2^ = 0.1, LASSO) and lung (*R*^2^ = 0.083, elastic net), respectively. Probabilistic finemapping of the marginal TWAS *Z* scores in the *VDAC2* region was then undertaken using a multi-tissue panel to assess evidence for whether *VDAC2* is the causal gene in this region. This locus was dense with genes, with 26 unique genes within the 90% credible set of causal genes. There was moderate evidence that *VDAC2* was the most probable causal gene at this locus given it had the largest absolute TWAS *Z*, the highest *PIP* for an individual model from the adrenal gland (*PIP* = 0.279), and a relatively large cumulative *PIP* for all 17 *VDAC2* models derived a variety of tissues that were finemapped (*PIP*_Cumulative_ = 0.683). Further investigation is thus needed to refine this locus.

There were a number of other genes that trended towards surviving multiple-testing correction in spleen and the other two tissues – including, *PLD4* in spleen and *STPG1* in lung. We subjected genes that trended towards multiple testing correction (two orders of magnitude above the Bonferroni threshold) to gene-set overrepresentation analysis to identify gene ontologies enriched for these genes, with *immune system process* and *immune response* overrepresented after multiple-testing correction, supporting the biological relevance of this signal (Supplementary Table 12). For instance, downregulation of tumour necrosis factor receptor gene *TNFRS19* in lung trended towards correlation with pneumonia, *Z* = -3.44, *P* = 5.78 × 10^−4^.

### Pneumonia displays genetic correlation with clinically significant phenotypes

We estimated genetic correlation between pneumonia and 180 GWAS using LDSR, with a significant non-zero estimate of genetic correlation obtained for twenty phenotypes after the application of multiple testing correction (*P* < 2.78 × 10^−4^, Supplementary Table 13, Fig. 2). Interestingly, the most significant genetic correlation was found between pneumonia and insomnia (*r*_g_ = 0.496, *SE* = 0.075, *P* = 3.55 × 10^−11^), which supports previous observational data that insomnia and reduced sleep duration increased the risk of developing pneumonia^48,49^. In addition, we uncovered significant genetic correlation with other clinically interesting phenotypes including forced vital capacity (*r*_g_ = -0.215, *SE* = 0.0361, *P* = 2.62 × 10^−9^), obesity (*r*_g_ = 0.3023, *SE* = 0.0466, *P* = 9.15 × 10^−11^), and HDL cholesterol (*r*_g_ = -0.2876, *SE* = 0.0664, *P* = 1.48 × 10^−5^).

**Figure 2.**
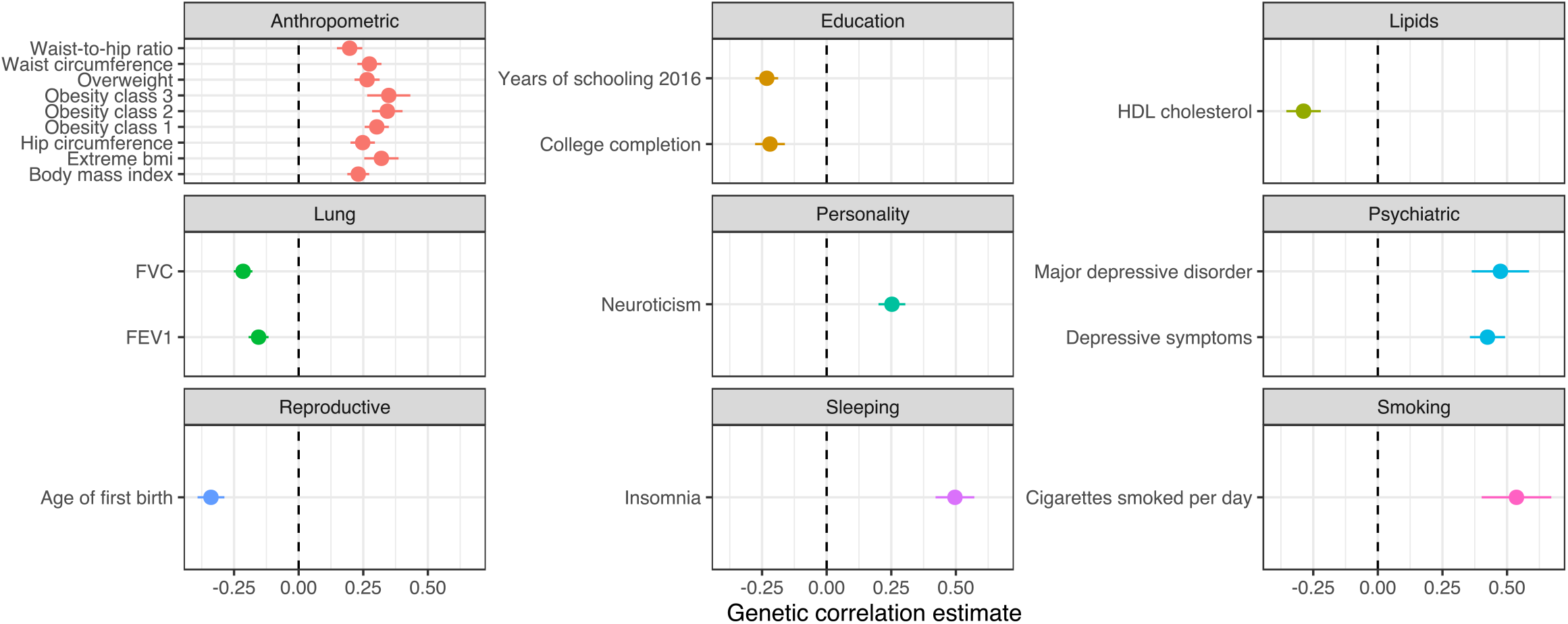
Estimates of genetic correlation between pneumonia and a panel of GWAS that survive multiple testing correction. Each panel of the forest plot is the estimate of genetic correlation by LD score regression (+/- its standard error, denoted by the error bars). The dotted lines represent a genetic correlation of zero. Panels were grouped by the phenotypic category of the trait subjected to LDSR.

A latent causal variable (LCV) model was then constructed for the most significantly correlated trait-pairs from each phenotypic category that comprised the 180 GWAS tested for genetic correlation. The LCV approach leverages the bivariate effect size distribution of SNPs in two GWAS and their LD scores to estimate a genetic causality proportion (GCP), such that, evidence of partial genetic causality can be distinguished from genetic correlation. We found strong evidence 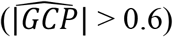 for partial genetic causality of cigarettes per day and HDL on pneumonia. Both posterior mean GCP estimates were significantly different from zero, however, the estimate of HDL → pneumonia 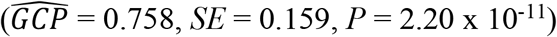 was more precise than that of cigarettes per day → pneumonia 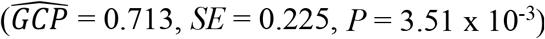. The magnitude of the potential causal relationship between HDL and pneumonia was further investigated using mendelian randomisation (MR). Given the biological overlap between the genetic architecture of HDL and other lipid classes, we constructed a multivariable MR model that conditioned HDL instrumental variables on their association with LDL cholesterol and triglycerides, obtaining the SNP-exposure estimates from the Willer *et al*. global lipids genetics consortium paper, as has been outlined elsewhere ^29,50^. There was no evidence of a causal effect of HDL (*P* = 0.65) or LDL (*P* = 0.47) conditioned on the remaining lipid classes, although there was nominal evidence of a risk increasing effect of triglycerides on pneumonia – *β* = 0.058, *SE* = 0.028, *P* = 0.035. These data highlight the complexities of distinguishing between confounding pleiotropy and evidence for causal relationships, with further work needed to resolve whether the directionally disproportionate variant effect sizes for HDL → pneumonia captured by the LCV model represent a true evidence for a causal relationship or whether other factors like triglycerides may explain this relationship. In addition, the effect of other confounders like BMI on these relationships cannot also be ruled out, although BMI did not show evidence of a causal effect in the LCV model.

### Opportunities for drug repurposing by leveraging the genetic architecture of pneumonia

We sought to interrogate the pneumonia GWAS to propose novel drug repurposing candidates that could be useful to treat patients diagnosed with pneumonia more effectively. Firstly, we utilised a very liberal approach which identified genes that were targeted by approved drugs outside the MHC region physically mapped to loci associated with pneumonia at a minimum of a suggestive significance threshold (*P* < 1 × 10^−5^). There were five such genes that displayed a high confidence interaction with an approved pharmacological agent with a known mechanism of action – *IL6R, SCNN1A, ATP2B1, ERBB2*, and *STAT5B*). For instance, Tocilizumab is a monoclonal antibody that targets *IL6R* which has been suggested as a repurposing opportunity to use for severe illness following SARS-CoV2 infection, although results from randomised controlled trials have been mixed in terms of efficacy^51^. We examined these genes in the TWAS analyses, however, only *IL6R* was significantly *cis*-heritable in one of the three tissues we utilised. Interestingly, there was a trend towards a correlation between downregulation of *IL6R* and increased odds of pneumonia, which would not support the use of anti-IL-6 receptor agents like tocilizumab – *Z* = -3.148, *P* = 1.64 × 10^−3^. In contrast, the lead SNP and odds increasing allele of the *IL6R* locus in this GWAS has been associated with increased *IL6R* levels in a protein quantitative trait loci study (pQTL), which would support the efficacy of tocilizumab. We caution that the pQTL signal is in high LD with a missense variant (rs2228145), and thus, antigen binding affinity may be altered to create an artefactual pQTL association. These antigen-binding affinity related effects require further investigation, particularly in light of the phenomenon of the non-synonymous rs2228415 C allele displaying correlation with increased protein abundance from the pQTL study^52^ but with decreased expression via RNAseq derived eQTL estimates from whole blood by GTEx, along with decreased CRP levels in the UK biobank, a well-characterised biomarker of IL-6 receptor (IL-6R) inhibition. Previous functional analyses of the rs2228145 non-synonymous allele have demonstrated that it likely impairs IL6 signalling, with increased expression of soluble circulating IL-6R but downregulation of the membrane bound isoform^53^. As a result, we conclude that based on the genetic association alone that the anti-inflammatory effect of IL-6R blockade may have a risk-increasing impact on pneumonia, although further work is required to evaluate tocilizumab as a potential repurposing opportunity, particularly as its efficacy would likely be dependent on its temporal application in the clinical course of the disease.

A panel of 50 metabolites and blood cell count phenotypes from the UK biobank (UKBB) with moderate to high confidence SNP-based heritability estimates were then tested for genetic correlation with pneumonia (Supplementary Table 14). The concept underlying this is that if there is evidence of a relationship between a biochemical trait and pneumonia, then a drug which modulates that trait in a risk-decreasing direction could be clinically useful, and thus, repurposed for pneumonia. We found 13 biochemical traits from the UKBB panel that were correlated with pneumonia after multiple-testing correction (Supplementary Table 15). The most significant correlation was a positive genetic correlation with triglycerides, followed by a positive correlation with glycaeted haemoglobin (HbA1c), which is interesting given previous evidence that hyperglycaemia has a deleterious effect on lung function^27,54^. The correlation between HbA1c and pneumonia may not be necessarily driven by glycaemic biology, particularly as HbA1c is strongly influenced by haematological factors, although we did observe weak evidence for a positive correlation between glucose and pneumonia that does not survive multiple testing correction (*P* = 0.01, Supplementary Table 15). It should be noted that this UKBB sample is a larger sample size than the triglyceride GWAS subjected to LDSR in the broad LDhub GWAS panel from earlier in the manuscript, and thus, the estimate is more significant in this analysis. In addition, the HDL GWAS from the UKBB was not included due to anomalous estimates of large standard error when considering its heritability estimate (Supplementary Methods). There was only very weak evidence from the LCV model for a potential causal influence of triglycerides on pneumonia (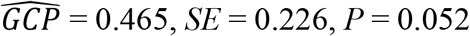, Figure 3a), although this broadly supports the results of the lipid multivariable MR described earlier in the manuscript that provided nominal evidence of a deleterious impact of increased triglycerides. We attempted to replicate the earlier multivariable MR results using the LDL, HDL, and triglyceride GWAS from the UK biobank as the exposure traits rather than the Willer *et al*. global lipids genetics consortium GWAS, with an analogous result that demonstrated an odds-increasing effect of triglycerides but a non-significant effect of LDL or HDL cholesterol (Supplementary Table 16). We followed this up by estimating a univariable estimate of triglycerides → pneumonia, with a 5.4% mean increase in the odds of pneumonia per standard deviation increase in triglycerides across the five MR methods utilised (Supplementary Methods, Supplementary Table 17). The causal estimate was only statistically significant in the case of the two IVW estimators: OR = 1.056 [95% CI: 1.01, 1.104], *P* = 0.017 (IVW multiplicative random effects), however, the remaining models had relatively consistent point estimates of the effect of triglycerides on pneumonia (Figure 3b). There was some nominal evidence of heterogeneity amongst IV exposure-outcome estimates (*Q* = 212.51, *df* = 168, *P* = 0.01), however, the Egger intercept was not significantly different from – thus, we observed no direct statistical evidence of confounding pleiotropy. As a result, these data suggest a potential repurposing opportunity for drugs prescribed for hypertriglyceridemia, such as statins and fibrates, with the caveat that the MR and LCV models from this study provide only relatively weak to moderate support, particularly as the mean posterior GCP estimate was low, and thus, genetic correlation could contaminate the MR estimate.

**Figure 3.**
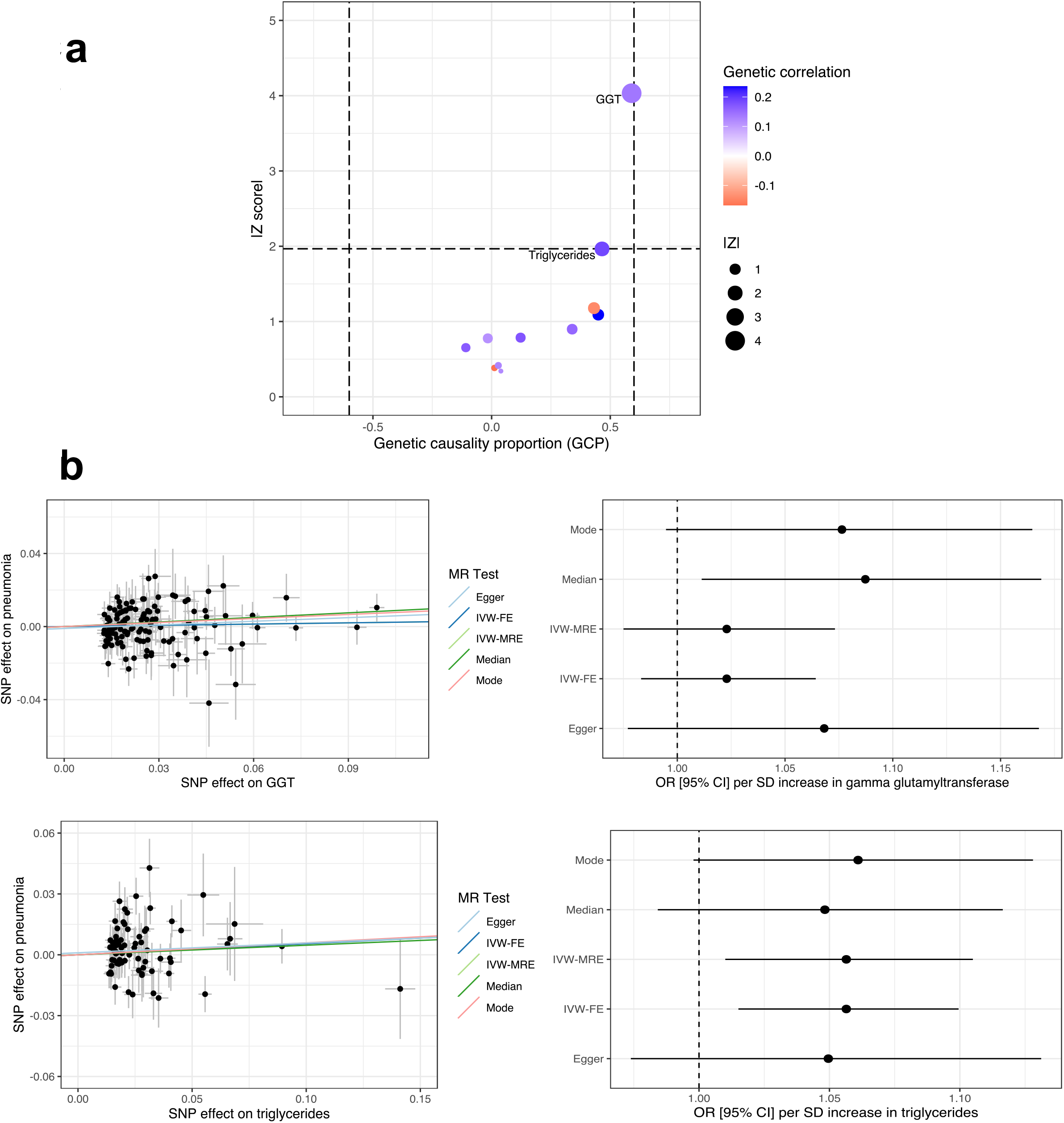
Investigating the potential utility of modulating biochemical traits as drug repurposing opportunities for pneumonia. (**a**) Latent causal variable models constructed between genetically correlated trait pairs after Bonferroni correction. Each point represents the genetic causality proportion, with the *y* axis denoting the precision of the GCP estimate, that is, its *Z* score. The genetic correlation estimate between the two traits was utilised to shade the points, with the larger points also indicative of a larger GCP *Z* score. A positive GCP estimate is indicative of partial genetic causality of the biochemical trait → pneumonia, whilst a negative estimate represents the converse. (**b**) A Mendelian randomisation (MR) analysis of the effect of genetically proxied gamma glutamyl-transferase (GGT) and triglycerides on the odds of pneumonia. The scatter plot visualises the effect of each instrumental variable SNP on GGT or triglycerides verses its effect on pneumonia, with the regression trend line the MR estimate from each of the five models implemented. Similarly, the forest plot indicates the pneumonia odds ratio for each of the MR models, with the error bar indicative of the 95% confidence intervals. The MR models were as follows: mode = weighted mode estimator, median = weighted median estimator, IVW-FE = inverse-variance weighted estimator with fixed effects, IVW-MRE = inverse-variance weighted estimator with multiplicative random effects, Egger = MR-Egger regression.

The remaining LCV models for biochemical traits genetically correlated with pneumonia did not indicate any strong evidence of partial genetic causality, with the exception of a putative effect of gamma-glutamyltransferase (GGT) on pneumonia that approximately reached the threshold for a strong point estimate of the 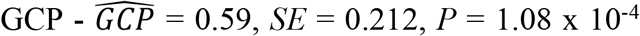. GGT is an enzyme that is commonly characterised as a biomarker of liver dysfunction, with some evidence of an immunological role for this enzyme, as well as its involvement in alveolar gas exchange. We further investigated this putative causal relationship by leveraging 207 independent SNPs associated with GGT (*P* < 5 × 10^−8^) in the UK biobank as IVs – approximated variance explained by IVs = 6.47%, *F*-statistic = 114.91. The five MR models implemented in this study yielded a roughly similar effect size per SD increase in GGT that increased the odds of pneumonia (mean pneumonia OR per SD GGT increase = 1.055, Supplementary Table 18). However, the estimates were not statistically significant, with the exception of the weighted median model (OR = 1.087 [95% CI: 1.01, 1.17], *P* = 0.025). There was evidence of heterogeneity amongst the IV exposure-outcome effects, which may be indicative of confounding pleiotropy (*Q* = 301.87, *df* = 206, *P* = 1.501 × 10^−5^), although given the large number of IVs observed heterogeneity is not a surprising phenomenon. Importantly, there was no statistical evidence of pleiotropy from the intercept of the MR egger model. We caution that statistical approaches to assess pleiotropy do not and cannot rule out a confounding influence on the MR estimate, and detailed biological annotation of the IVs would be warranted to investigate this further. In summary, there may be a causal relationship between GGT and increased odds of pneumonia which could support inhibition of this enzyme as a treatment target, with several GGT inhibitors under active development and explored for use in respiratory illness^55,56^.

Furthermore, we compared the MR estimate of the effect of a standard deviation increase in GGT and triglyceride concentration on the odds of pneumonia to an observational estimate from the UK biobank sample (Supplementary Methods). We found that the observational association between a standard deviation increase in triglyceride concentration and pneumonia was analogous to the univariable and multivariable MR estimates (OR = 1.059 [95% CI: 1.042, 1.076], *P* = 2 × 10^−12^), whilst the observed association between GGT and pneumonia was stronger than the MR estimate, with each standard deviation associated with a 13.54% [95% CI: 12.43%, 14.56%] increase in the odds of pneumonia amongst UK biobank participants. Interestingly, these associations were also consistent amongst individuals in the UK biobank with relatively lower risk of pneumonia, that is, non-smoking females aged 45 or younger at time of assessment – triglycerides: OR = 1.335 [95% CI: 1.13, 1.555], *P* = 2.67 × 10^−4^, GGT: OR = 1.354 [95% CI: 1.161, 1.553], *P* = 2.75 × 10^−5^. These variables are extremely heterogeneous and there are many potential confounders of the observed effect sizes, however, it supports the inferred relationship from the LCV and MR models.

### Precision drug repurposing to treat pneumonia

We implemented the *pharmagenic enrichment score* (PES) approach to identify drug repurposing candidates that could be targeted more precisely based on genetic risk^27,37^. Briefly, the PES is a genetic risk score specifically within a biological pathway that is targeted by approved drugs. The concept underlying the PES is that individuals with elevated genetic risk within a particular druggable set of genes may benefit from a pharmacological agent that modulates the pathway in question. Firstly, we identified five druggable pathways that displayed an enrichment of the common variant genetic architecture of pneumonia at one of four *P*-value thresholds for the inclusion of variants in the model (FDR < 0.05, Table 2, Supplementary Table 19, Supplementary Methods). These included two complement-related pathways, *p53 signalling*, and *bile acid metabolism*.

**Table 2.** Candidate druggable gene-sets that could be utilised to calculate *pharmagenic enrichment scores*.

| PES gene-set | $P_T^1$ | $P$ | Example drug/nutraceutical |
| --- | --- | --- | --- |
| P53 signalling pathway | 0.05 | $2.13 \times 10^{-8}$ | Fostamatinib |
| Lectin induced complement pathway | 0.05 | $2.98 \times 10^{-8}$ | Human immunoglobulin |
| Complement pathway | 0.05 | $6.81 \times 10^{-8}$ | Zinc |
| Bile acid metabolism | 0.005 | $1 \times 10^{-6}$ | Atorvastatin |
| RIG-I-like receptor signalling pathway | 0.005 | $1.58 \times 10^{-5}$ | Etanercept |

<sup>1</sup> $P_T = P$ value threshold for variant inclusion in the model

There were a number of diverse compounds that targeted these pathways, with compounds identified as repurposing candidates through testing whether there was a statistically significant overrepresentation of their targets in the gene-set, along with high confidence single drug-gene interactions (Supplementary Tables 20,21). For instance, targets of the micronutrient zinc were overrepresented amongst genes in the *Complement pathway* gene-set, supporting previous evidence that zinc can modulate complement activation ^57,58^.

#### Characteristics of pneumonia pharmagenic enrichment scores

We investigated the properties of these five scores in the UKBB cohort. Interestingly, there were no large correlations (all *r* < 0.07) between the PES and a genome-wide polygenic risk score for pneumonia, which supports our hypothesis that pathway-based risk scores may provide novel biological insights that are not encompassed by PRS constructed from variants throughout the genome. Both the genome wide PRS and the PES profiles were not significantly associated with pneumonia diagnosis (self-reported/clinically ascertained or clinically ascertained only, Supplementary Tables 22,23, Supplementary Methods). This is perhaps not surprising given the low SNP heritability of pneumonia and the heterogeneity of the phenotype – however, we believe this does not preclude the relevance of PES at an individual level. For instance, given the putative relationship between hypertriglyceridemia and increased odds of pneumonia, as supported by this study, we tested the relationship between the pneumonia *Bile acid metabolism* PES and measured triglycerides in the UKBB. There was a small but significant positive correlation between the PES and triglyceride concentration – *β* = 0.01, *SE* = 0.002, *P* = 1.69 × 10^−4^, which was significant even after adjusting for statin use or using triglyceride values as the outcome variable winsorized at three standard deviations above the mean to guard against an excessive influence of outliers (*β* = 0.01, *SE* = 0.002, *P* = 1.43 × 10^−4^). This relationship with triglycerides was not seen using genome wide PRS (*β* = 1.57 × 10^−4^, *SE* = 0.002, *P* = 0.93).

#### Associations between pneumonia pharmagenic enrichment scores and host susceptibility to infection

The relationship between PES, as a function of host genetic susceptibility to pneumonia, and antibody response to infection was then assessed in a subset of the UKBB with these data available and passing our genotyping QC thresholds (N = 6443). We implemented these analyses given the host-immune response is directly relevant to pneumonia and if the PES displays distinct correlations with antibody response compared to genome-wide pneumonia PRS, it would emphasise the potential biological utility of the PES framework. Firstly, we tested the association between each PES and antibody response to 28 antigens amongst individuals with detectable levels of antibodies for 14 infections (seroreactivity, Fig. 4c, Supplementary Table 24). The strongest relationship between a PES profile and seroreactivity was a positive correlation between the *Complement pathway* PES and IgG response to the major capsid protein VP1 of the BK polyomavirus, with each SD increase in the PES associated with a 0.05 (0.01) SD increase in antibody response, *P* = 4.1 × 10^−4^, which trended towards surviving multiple testing correction (*q* = 0.07). There was no association between genome wide PRS and IgG mediated response to this antigen. There were other nominally significant correlations observed (Fig. 4a), with most of these correlations positive and potentially indicative of an increased immune response. In addition, susceptibility to infection (seropositivity) was also investigated (Supplementary Table 25). For instance, a nominal association uncovered between *RIG-I-like receptor signalling pathway* and decreased odds of herpes simplex virus-1 infection (OR = 0.93 per SD in score [95% CI: 0.87, 0.98], *P* = 4 × 10^−3^), whilst *Complement pathway* PES was nominally associated with increased odds of positive *H. pylori* serostatus - OR = 1.07 per SD in score [95% CI: 1.01, 1.12], *P* = 0.02.

**Figure 4.**
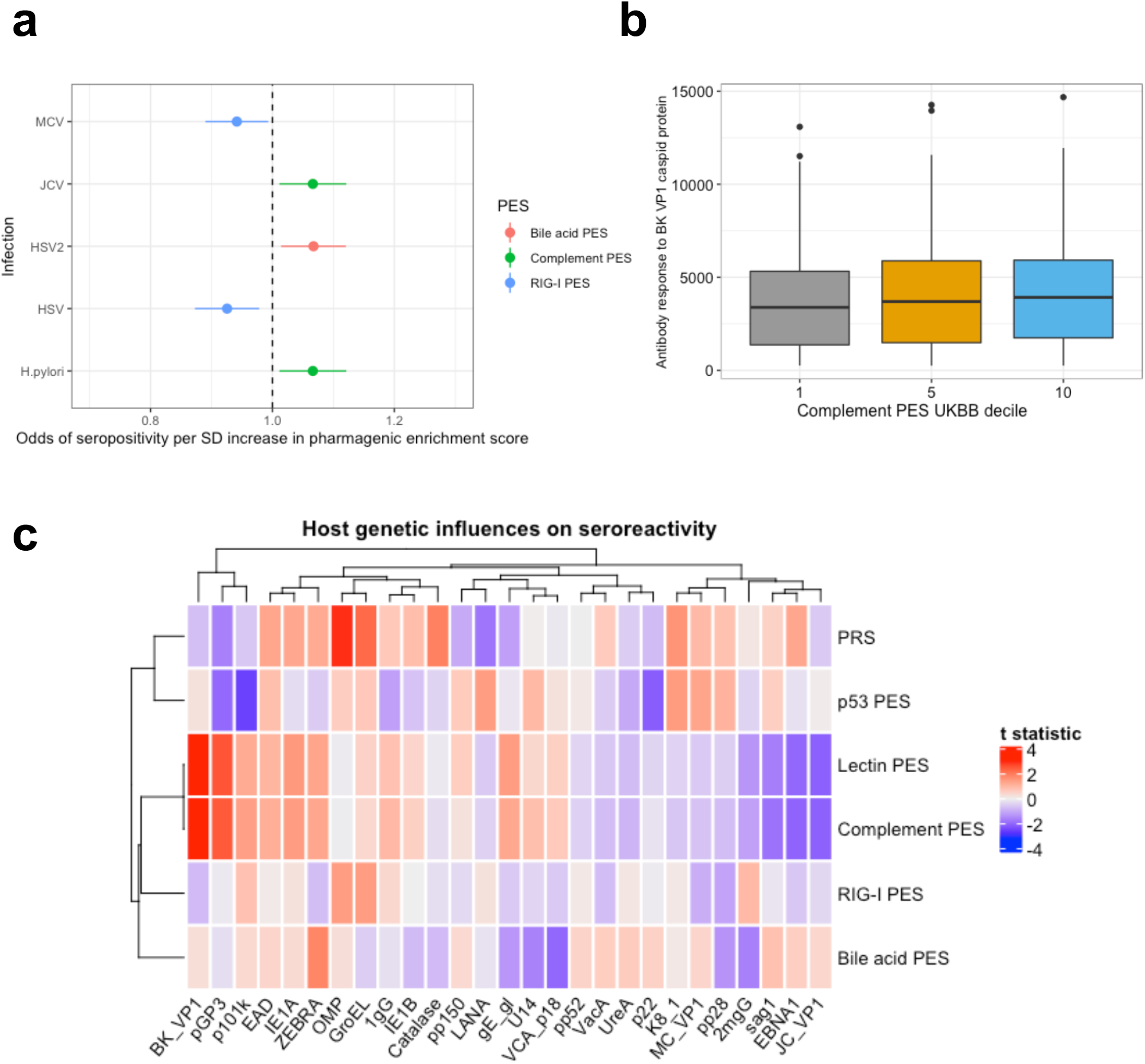
The relationship between host genetics pneumonia pathway based *pharmagenic enrichment scores* and antibody response. (**a**) Nominally significant associations between pneumonia PES and seropositivity as a binary variable. The forest plot denotes the odds ratio of each infection serostatus (error bars represent 95% confidence intervals) per SD increase in the PES tested. The infections abbreviated are as follows: MCV = Merkel Cell Polyomavirus, JCV = Human Polyomavirus JCV, HSV = herpes simplex virus 1, HSV2 = herpes simplex virus 2, *H. pylori* = Helicobacter pylori. (**b**) IgG response (mean fluorescence intensity) to the major capsid protein VP1 of the BK polyomavirus in the 10^th^ percentile (1^st^ decile, denoted 1), 50^th^ percentile (5^th^ decile, denoted 5), and 90^th^ percentile (10^th^ decile, denoted 10) of the complement pathway PES amongst the genotyped subset of the UKBB subjected to antibody screening included in our analyses. (**c**) Heatmap of the regression *t* statistic (beta/standard error) for the correlation between each PES and PRS with respective IgG response to antigens amongst individuals seropositive for that infection. Hierarchical clustering was applied to the rows and columns using Pearson’s correlation distance.

## DISCUSSION

In this study, we uncovered the first significant association signals for lifetime pneumonia susceptibility outside of the MHC region. Interestingly, there was also a novel low frequency variant in the MHC itself which reached genome-wide significant that confers a relatively large (∼ 59%) increase in the odds of pneumonia, reflecting the immense heterogeneity spanned within this region. Further analyses of the MHC signal are warranted, particularly to deconvolve specific HLA types that may contribute to pneumonia susceptibility and progression. Each of the loci beyond the MHC identified this study were relatively complex, although we were able to derive a 95% credible set for the most significant non-MHC locus on chromosome 11 that implicated the mucin gene *MUC5AC*. It should be noted that our fine-mapping approach is relatively biologically naïve as it assumes a single causal variant, and thus, the involvement of other genes remains unclear. *MUC5AC* is an interesting candidate given that it has been previously implicated in the pathogenesis of respiratory illness and the role of mucins in physical defence against pathogens via mucociliary clearance^59^. This heavily glycosylated protein is lowly expressed in normal respiratory epithelium, however, is upregulated upon in response to perturbagens, such as viral infection^60,61^. We posit that upregulation of *MUC5AC* may be deleterious in the context of pneumonia given recent evidence that this protein can enhance airway inflammation induced by viral infection^62^, although dissection of the mechanisms of variants in this locus are warranted, particularly given the apparent discordant relationship of this signal we observed on the odds on asthma. Interestingly, there are some preliminary data that suggests *MUC5AC* is upregulated in the airway mucus of patients with severe COVID-19, although these studies were conducted using small sample sizes^63,64^. There was also evidence of a more expansive polygenic signal amongst mucin and beta-galactoside/N-acetylgalactosaminide genes in the *termination of O-glycan biosynthesis pathway*, whereby sialic acid residues conjugated to mucins can terminate O-glycan biosynthesis^65^. Interestingly, therapies specifically targeting mucin-linked O-glycosylation are now under active development, including a recently proposed hexosamine analog that demonstrated potent inhibition of O-glycan biosynthesis and downregulation of neutrophil infiltration in rodents^66^.

Drug repurposing is an attractive downstream application for GWAS, and we demonstrate its potential utility for pneumonia through three distinct methods. Causal inference between biochemical traits and pneumonia may provide repositioning opportunities along with a greater understanding of pathological mechanisms in the disorder. For instance, we reveal evidence to suggest a potential protective effect of HDL cholesterol and a deleterious impact of elevated triglycerides on the odds of pneumonia. These data support observational data that lower baseline HDL and elevated triglycerides have risk-increasing properties for pneumonia^67–69^,. We emphasise that our data only provided weak to moderate support for a causal influence of lipid abundance on pneumonia, and replicated, well-powered randomised controlled trials are needed to definitively assess the suitability of lipid-modifying agents like statins. A key limitation of proposing drug repurposing candidates for the phenotype as a singular entity is that it ignores the inherent heterogeneity of pneumonia onset and clinical course. It has been shown previously that individual genes supported by GWAS significantly increase the likelihood of a candidate drug being successfully approved from the phase I stage ^70^, however, the relevance of more expansive association signals relating to biological networks for precision medicine remains unclear As a result, we sought to implement an approach which seeks to target drug repositioning opportunities to those with elevated genetic risk within pathways relevant to the compound (PES, one of the prioritised pathways for the construction of a PES was the *Bile acid metabolism* gene-set, further supporting the relevance of compounds that modulate cholesterol. In the UKBB cohort, we demonstrated that these scores were distinct from genome-wide PRS, and thus, may offer biological insights that were missed by using a genome-wide score. For example, the *Bile acid metabolism* PES was positively correlated with triglyceride levels, whilst this was not observed for PRS. We caution that one cannot draw a causal inference from this relationship. In addition, the putative relationship between PES and IgG response to antigens is biologically relevant both in terms of increased and decreased antibody response. One can conceptualise the phenotype of pneumonia as having a contribution from increased likelihood of infection, but also an aberrant inflammatory response once infected with a pathogen. The distinct signal observed with these IgG phenotypes for the pathway-based PES compared to genome-wide PRS, therefore, further highlights the potential utility of the PES framework. Further work is required to evaluate the suitability of compounds that modulate the PES pathways of interest and to categorise drug repurposing candidates based on their suitability for prevention and/or treatment of pneumonia. The key advantage of the PES framework is that only individuals with relevant genetic background in a pathway of interest would be prioritised for the respective repurposing candidate, which would be useful given the polygenic nature of complex disorders. Detailed discussion of the strengths and limitations of the PES methodology have been featured in previous publications^27,37^.

There are a number of important limitations that should be considered in light of the pneumonia GWAS itself and our drug repurposing analyses. Firstly, this GWAS was conducted using samples from European ancestry as large, diverse, genotyped cohorts with pneumonia status information are not yet available. It will be critical to translate findings related to host-genetic influences on pneumonia that future efforts strive to collect trans-ancestral data, particularly due to concerns about the portability of European GWAS signals and the advantages in finemapping afforded by including multiple ancestries^71^. The SNP heritability for pneumonia derived in this study was also relatively low, and it remains unclear how heterogeneity amongst the phenotype definition of pneumonia may contribute to this. In other words, given that pneumonia is caused by a variety of factors and may go undiagnosed in some individuals, detailed phenotyping data would potentially assist in resolving the genetic architecture of this disorder. For example, a GWAS on susceptibility verses pneumonia severity will likely reveal different biological insights. This could also be aided by stratified analyses by age, given pneumonia is more pervasive in the elderly. The putative drug repurposing candidates suggested in this study must also be viewed in light of the low heritability of pneumonia and need for clinical validation. Despite these challenges, we believe that further resolving the host-genetic architecture of pneumonia will be invaluable to public health efforts to more effectively prevent and manage the illness. In summary, we revealed novel genome-wide significant loci associated with life-time pneumonia susceptibility beyond the MHC region. These data provided some support for the potential utility of triglycerides and GGT as treatment targets for pneumonia, however, randomised controlled trials are now required to establish the efficacy of such interventions. Moreover, the *pharmagenic enrichment score* approach may provide a precision medicine-based intervention for drug repurposing for pneumonia prophylaxis and treatment given an individual’s composition of genetic risk. The properties of these scores and the prospects of integrating them with other clinical metrics warrants further research.

## Supporting information

Supplementary Methods and Results

## Data Availability

In accordance with the 23andMe data sharing policies, we are only able to release 10,000 SNPs from our meta-analysis, which we have made available on GitHub (https://github.com/Williamreay/Pneumonia_meta_GWAS_drug_repurposing/tree/master/Summary_statistics). The full GWAS summary statistics for the 23andMe discovery data set will be made available through 23andMe to qualified researchers under an agreement with 23andMe that protects the privacy of the 23andMe participants.
Researchers wishing to recapitulate our meta-analysis can apply for access for the 23andMe subset of the study (https://research.23andme.com/dataset-access/), and then meta-analyse with FinnGen release 3 summary statistics as described in our manuscript. Code utilised in this study is available also on GitHub - https://github.com/Williamreay/Pneumonia_meta_GWAS_drug_repurposing.

## ACKNOWLEDGEMENTS

We wish to acknowledge the participants and investigators of FinnGen study and the 23andMe Inc. study from which these data were derived for the GWAS meta-analysis. In addition, his research has been conducted using the UK Biobank Resource under the application 58432. This study was supported by an NHMRC project grant (1147644). M.J.C. is supported by an NHMRC Senior Research Fellowship (1121474).

## CONSORTIA

### The 23andMe Research Team

Michelle Agee^3^, Babak Alipanahi^3^, Robert K. Bell^3^, Katarzyna Bryc^3^, Sarah L. Elson^3^, Pierre Fontanillas^3^, Nicholas A. Furlotte^3^, Barry Hicks^3^, David A. Hinds^3^, Karen E. Huber^3^, Ethan M. Jewett^3^, Yunxuan Jiang^3^, Aaron Kleinman^3^, Keng-Han Lin^3^, Nadia K. Litterman^3^, Jennifer C. McCreight^3^, Matthew H. McIntyre^3^, Kimberly F. McManus^3^, Joanna L. Mountain^3^, Elizabeth S. Noblin^3^, Carrie A. M Northover^3^, Steven J. Pitts^3^, G. David Poznik^3^, J. Fah Sathirapongsasuti^3^, Janie F. Shelton^3^, Suyash Shringarpure^3^, Chao Tian^3^, Joyce Y. Tung^3^, Vladimir Vacic^3^, Xin Wang^3^ & Catherine H. Wilson^3^.

## AUTHOR INFORMATION

**School of Biomedical Sciences and Pharmacy, Faculty of Health and Medicine, The University of Newcastle, Callaghan, NSW, 2308, Australia**

William R. Reay, Michael P. Geaghan, and Murray J. Cairns

**Hunter Medical Research Institute, Newcastle, NSW, 2305, Australia**

William R. Reay, Michael P. Geaghan, and Murray J. Cairns

**23andMe Inc**., **Sunnyvale, CA, 94086, United States of America**

The 23andMe Research Team

## WEB RESOURCES

DGIdb v4.2.0 - https://www.dgidb.org/

FAVOR - http://favor.genohub.org/

FOCUS version July 24 2019 - https://github.com/bogdanlab/focus

FUMA v1.3.6 - https://fuma.ctglab.nl/

FUSION version May 29 2020 - https://github.com/gusevlab/fusion_twas

LCV version March 15 2019 - https://github.com/lukejoconnor/LCV

LDSR v1.0.1 - https://github.com/bulik/ldsc

MAGMA v.1.07b - https://ctg.cncr.nl/software/magma

METAL version March 2011 - https://genome.sph.umich.edu/wiki/METAL_Quick_Start

MR-PRESSO v1 - https://github.com/rondolab/MR-PRESSO/commits/master

mtCOJO (GCTA version 1.93.2 beta mac) –

https://cnsgenomics.com/software/gcta/#mtCOJO

PLINK2 v.2.00a3LM - https://www.cog-genomics.org/plink/2.0/

PRSice-2 v.2.3.3 (linux) - https://www.prsice.info/

TwoSampleMR v.0.5.5 - https://github.com/MRCIEU/TwoSampleMR

VEP GRCh37 release 101 August 2020 –

https://grch37.ensembl.org/Homo_sapiens/Tools/VEP?db=core

WebGestaltR v.4.0.2 - https://github.com/bzhanglab/WebGestaltR

## DATA AND CODE AVAILABILITY

All data in this study are publicly available, summary statistics from 23andMe Inc. can be obtained upon application to the company. As per 23andMe data sharing policies, we are only able to release 10,000 SNPs from our meta-analysis, which we have made available on GitHub (https://github.com/Williamreay/Pneumonia_meta_GWAS_drug_repurposing/tree/master/Summary_statistics). The full GWAS summary statistics for the 23andMe discovery data set will be made available through 23andMe to qualified researchers under an agreement with 23andMe that protects the privacy of the 23andMe participants.

Researchers wishing to recapitulate our meta-analysis can apply for access for the 23andMe subset of the study (https://research.23andme.com/dataset-access/), and then meta-analyse with FinnGen release 3 summary statistics as described in our manuscript. Code utilised in this study is available also on GitHub - https://github.com/Williamreay/Pneumonia_meta_GWAS_drug_repurposing.

## REFERENCES

1. Mackenzie, G. (2016). The definition and classification of pneumonia. Pneumonia (Nathan) 8, 14.

2. Restrepo, M.I., Faverio, P., and Anzueto, A. (2013). Long-term prognosis in community-acquired pneumonia. Curr Opin Infect Dis 26, 151–158.

3. McAllister, D.A., Liu, L., Shi, T., Chu, Y., Reed, C., Burrows, J., Adeloye, D., Rudan, I., Black, R.E., Campbell, H., et al. (2019). Global, regional, and national estimates of pneumonia morbidity and mortality in children younger than 5 years between 2000 and 2015: a systematic analysis. Lancet Glob Health 7, e47–e57.

4. Obel, N., Christensen, K., Petersen, I., Sørensen, T.I.A., and Skytthe, A. (2010). Genetic and Environmental Influences on Risk of Death due to Infections Assessed in Danish Twins, 1943–2001. American Journal of Epidemiology 171, 1007–1013.

5. Tian, C., Hromatka, B.S., Kiefer, A.K., Eriksson, N., Noble, S.M., Tung, J.Y., and Hinds, D.A. (2017). Genome-wide association and HLA region fine-mapping studies identify susceptibility loci for multiple common infections. Nat Commun 8, 599.

6. Willer, C.J., Li, Y., and Abecasis, G.R. (2010). METAL: fast and efficient meta-analysis of genomewide association scans. Bioinformatics 26, 2190–2191.

7. Watanabe, K., Taskesen, E., van Bochoven, A., and Posthuma, D. (2017). Functional mapping and annotation of genetic associations with FUMA. Nat Commun 8, 1826.

8. Zhu, Z., Zheng, Z., Zhang, F., Wu, Y., Trzaskowski, M., Maier, R., Robinson, M.R., McGrath, J.J., Visscher, P.M., Wray, N.R., et al. (2018). Causal associations between risk factors and common diseases inferred from GWAS summary data. Nat Commun 9, 224.

9. Liu, M., Jiang, Y., Wedow, R., Li, Y., Brazel, D.M., Chen, F., Datta, G., Davila-Velderrain, J., McGuire, D., Tian, C., et al. (2019). Association studies of up to 1.2 million individuals yield new insights into the genetic etiology of tobacco and alcohol use. Nat Genet 51, 237–244.

10. Ferreira, M.A.R., Mathur, R., Vonk, J.M., Szwajda, A., Brumpton, B., Granell, R., Brew, B.K., Ullemar, V., Lu, Y., Jiang, Y., et al. (2019). Genetic Architectures of Childhood- and Adult-Onset Asthma Are Partly Distinct. Am J Hum Genet 104, 665–684.

11. Giambartolomei, C., Vukcevic, D., Schadt, E.E., Franke, L., Hingorani, A.D., Wallace, C., and Plagnol, V. (2014). Bayesian test for colocalisation between pairs of genetic association studies using summary statistics. PLoS Genet. 10, e1004383.

12. Zhou, W., Nielsen, J.B., Fritsche, L.G., Dey, R., Gabrielsen, M.E., Wolford, B.N., LeFaive, J., VandeHaar, P., Gagliano, S.A., Gifford, A., et al. (2018). Efficiently controlling for case-control imbalance and sample relatedness in large-scale genetic association studies. Nat. Genet. 50, 1335–1341.

13. Bulik-Sullivan, B., Finucane, H.K., Anttila, V., Gusev, A., Day, F.R., Loh, P.-R., ReproGen Consortium, Psychiatric Genomics Consortium, Genetic Consortium for Anorexia Nervosa of the Wellcome Trust Case Control Consortium 3, Duncan, L., et al. (2015). An atlas of genetic correlations across human diseases and traits. Nat. Genet. 47, 1236–1241.

14. Wellcome Trust Case Control Consortium, Maller, J.B., McVean, G., Byrnes, J., Vukcevic, D., Palin, K., Su, Z., Howson, J.M.M., Auton, A., Myers, S., et al. (2012). Bayesian refinement of association signals for 14 loci in 3 common diseases. Nat. Genet. 44, 1294–1301.

15. Benner, C., Havulinna, A.S., Järvelin, M.-R., Salomaa, V., Ripatti, S., and Pirinen, M. (2017). Prospects of Fine-Mapping Trait-Associated Genomic Regions by Using Summary Statistics from Genome-wide Association Studies. Am J Hum Genet 101, 539–551.

16. de Leeuw, C.A., Mooij, J.M., Heskes, T., and Posthuma, D. (2015). MAGMA: generalized gene-set analysis of GWAS data. PLoS Comput. Biol. 11, e1004219.

17. Liberzon, A., Birger, C., Thorvaldsdóttir, H., Ghandi, M., Mesirov, J.P., and Tamayo, P. (2015). The Molecular Signatures Database (MSigDB) hallmark gene set collection. Cell Syst 1, 417–425.

18. Liu, Y., and Xie, J. (2020). Cauchy Combination Test: A Powerful Test With Analytic p - Value Calculation Under Arbitrary Dependency Structures. Journal of the American Statistical Association 115, 393–402.

19. Liu, Y., Chen, S., Li, Z., Morrison, A.C., Boerwinkle, E., and Lin, X. (2019). ACAT: A Fast and Powerful p Value Combination Method for Rare-Variant Analysis in Sequencing Studies. Am. J. Hum. Genet. 104, 410–421.

20. Turkmen, A., and Lin, S. (2017). Are rare variants really independent? Genet. Epidemiol. 41, 363–371.

21. Talluri, R., and Shete, S. (2013). A linkage disequilibrium-based approach to selecting disease-associated rare variants. PLoS ONE 8, e69226.

22. Gusev, A., Ko, A., Shi, H., Bhatia, G., Chung, W., Penninx, B.W.J.H., Jansen, R., de Geus, E.J.C., Boomsma, D.I., Wright, F.A., et al. (2016). Integrative approaches for large-scale transcriptome-wide association studies. Nat. Genet. 48, 245–252.

23. Mancuso, N., Freund, M.K., Johnson, R., Shi, H., Kichaev, G., Gusev, A., and Pasaniuc, B. (2019). Probabilistic fine-mapping of transcriptome-wide association studies. Nat. Genet. 51, 675–682.

24. Zheng, J., Erzurumluoglu, A.M., Elsworth, B.L., Kemp, J.P., Howe, L., Haycock, P.C., Hemani, G., Tansey, K., Laurin, C., Early Genetics and Lifecourse Epidemiology (EAGLE) Eczema Consortium, et al. (2017). LD Hub: a centralized database and web interface to perform LD score regression that maximizes the potential of summary level GWAS data for SNP heritability and genetic correlation analysis. Bioinformatics 33, 272–279.

25. O’Connor, L.J., and Price, A.L. (2018). Distinguishing genetic correlation from causation across 52 diseases and complex traits. Nat. Genet. 50, 1728–1734.

26. Reay, W.R., Kiltschewskij, D.J., Geaghan, M.P., Atkins, J.R., Carr, V.J., Green, M.J., and Cairns, M.J. (2021). Genetic estimates of correlation and causality between blood-based biomarkers and psychiatric disorders (Psychiatry and Clinical Psychology).

27. Reay, W.R., El Shair, S.I., Geaghan, M.P., Riveros, C., Holliday, E.G., McEvoy, M.A., Hancock, S., Peel, R., Scott, R.J., Attia, J.R., et al. (2021). Genetic association and causal inference converge on hyperglycaemia as a modifiable factor to improve lung function. ELife 10, e63115.

28. Hemani, G., Zheng, J., Elsworth, B., Wade, K.H., Haberland, V., Baird, D., Laurin, C., Burgess, S., Bowden, J., Langdon, R., et al. (2018). The MR-Base platform supports systematic causal inference across the human phenome. ELife 7, e34408.

29. Willer, C.J., Schmidt, E.M., Sengupta, S., Peloso, G.M., Gustafsson, S., Kanoni, S., Ganna, A., Chen, J., Buchkovich, M.L., Mora, S., et al. (2013). Discovery and refinement of loci associated with lipid levels. Nat Genet 45, 1274–1283.

30. Bowden, J., Davey Smith, G., Haycock, P.C., and Burgess, S. (2016). Consistent Estimation in Mendelian Randomization with Some Invalid Instruments Using a Weighted Median Estimator. Genet. Epidemiol. 40, 304–314.

31. Hartwig, F.P., Davey Smith, G., and Bowden, J. (2017). Robust inference in summary data Mendelian randomization via the zero modal pleiotropy assumption. Int J Epidemiol 46, 1985–1998.

32. Bowden, J., Davey Smith, G., and Burgess, S. (2015). Mendelian randomization with invalid instruments: effect estimation and bias detection through Egger regression. Int J Epidemiol 44, 512–525.

33. Verbanck, M., Chen, C.-Y., Neale, B., and Do, R. (2018). Detection of widespread horizontal pleiotropy in causal relationships inferred from Mendelian randomization between complex traits and diseases. Nat. Genet. 50, 693–698.

34. Thompson, S.G., and Sharp, S.J. (1999). Explaining heterogeneity in meta-analysis: a comparison of methods. Stat Med 18, 2693–2708.

35. Bowden, J., Del Greco M F., Minelli, C., Davey Smith, G., Sheehan, N., and Thompson, J. (2017). A framework for the investigation of pleiotropy in two-sample summary data Mendelian randomization. Stat Med 36, 1783–1802.

36. Reay, W.R., Shair, S.E., Geaghan, M.P., Riveros, C., Holiday, E.G., McEvoy, M.A., Hancock, S., Peel, R., Scott, R.J., Attia, J.R., et al. (2020). Genetically informed precision drug repurposing for lung function and implications for respiratory infection (Respiratory Medicine).

37. Reay, W.R., Atkins, J.R., Carr, V.J., Green, M.J., and Cairns, M.J. (2020). Pharmacological enrichment of polygenic risk for precision medicine in complex disorders. Sci Rep 10, 879.

38. Sudlow, C., Gallacher, J., Allen, N., Beral, V., Burton, P., Danesh, J., Downey, P., Elliott, P., Green, J., Landray, M., et al. (2015). UK biobank: an open access resource for identifying the causes of a wide range of complex diseases of middle and old age. PLoS Med 12, e1001779.

39. Bycroft, C., Freeman, C., Petkova, D., Band, G., Elliott, L.T., Sharp, K., Motyer, A., Vukcevic, D., Delaneau, O., O’Connell, J., et al. (2018). The UK Biobank resource with deep phenotyping and genomic data. Nature 562, 203–209.

40. Choi, S.W., and O’Reilly, P.F. (2019). PRSice-2: Polygenic Risk Score software for biobank-scale data. GigaScience 8, giz082.

41. Mentzer, A.J., Brenner, N., Allen, N., Littlejohns, T.J., Chong, A.Y., Cortes, A., Almond, R., Hill, M., Sheard, S., McVean, G., et al. (2019). Identification of host-pathogen-disease relationships using a scalable Multiplex Serology platform in UK Biobank (Infectious Diseases (except HIV/AIDS)).

42. Gu, Z., Eils, R., and Schlesner, M. (2016). Complex heatmaps reveal patterns and correlations in multidimensional genomic data. Bioinformatics 32, 2847–2849.

43. Linden, S.K., Sutton, P., Karlsson, N.G., Korolik, V., and McGuckin, M.A. (2008). Mucins in the mucosal barrier to infection. Mucosal Immunol 1, 183–197.

44. Chen, H.-H., Shaw, D.M., Petty, L.E., Graff, M., Bohlender, R.J., Polikowsky, H.G., Zhong, X., Kim, D., Buchanan, V.L., Preuss, M.H., et al. (2021). Host genetic effects in pneumonia. The American Journal of Human Genetics 108, 194–201.

45. Campos, A.I., Kho, P.F., Vazquez-Prada, K.X., García-Marín, L.M., Martin, N.G., Cuéllar-Partida, G., and Rentería, M.E. (2020). Genetic susceptibility to pneumonia: A GWAS meta-analysis between UK Biobank and FinnGen (Respiratory Medicine).

46. Kim, K., Park, S., Park, S.Y., Kim, G., Park, S.M., Cho, J.-W., Kim, D.H., Park, Y.M., Koh, Y.W., Kim, H.R., et al. (2020). Single-cell transcriptome analysis reveals TOX as a promoting factor for T cell exhaustion and a predictor for anti-PD-1 responses in human cancer. Genome Med 12, 22.

47. Sekine, T., Perez-Potti, A., Nguyen, S., Gorin, J.-B., Wu, V.H., Gostick, E., Llewellyn-Lacey, S., Hammer, Q., Falck-Jones, S., Vangeti, S., et al. (2020). TOX is expressed by exhausted and polyfunctional human effector memory CD8 ^+^ T cells. Sci. Immunol. 5, eaba7918.

48. Lin, C.-L., Liu, T.-C., Chung, C.-H., and Chien, W.-C. (2018). Risk of pneumonia in patients with insomnia: A nationwide population-based retrospective cohort study. J Infect Public Health 11, 270–274.

49. Patel, S.R., Malhotra, A., Gao, X., Hu, F.B., Neuman, M.I., and Fawzi, W.W. (2012). A prospective study of sleep duration and pneumonia risk in women. Sleep 35, 97–101.

50. Burgess, S., and Thompson, S.G. (2015). Multivariable Mendelian randomization: the use of pleiotropic genetic variants to estimate causal effects. Am J Epidemiol 181, 251–260.

51. Parr, J.B. (2020). Time to Reassess Tocilizumab’s Role in COVID-19 Pneumonia. JAMA Intern Med.

52. Folkersen, L., Fauman, E., Sabater-Lleal, M., Strawbridge, R.J., Frånberg, M., Sennblad, B., Baldassarre, D., Veglia, F., Humphries, S.E., Rauramaa, R., et al. (2017). Mapping of 79 loci for 83 plasma protein biomarkers in cardiovascular disease. PLoS Genet 13, e1006706.

53. Ferreira, R.C., Freitag, D.F., Cutler, A.J., Howson, J.M.M., Rainbow, D.B., Smyth, D.J., Kaptoge, S., Clarke, P., Boreham, C., Coulson, R.M., et al. (2013). Functional IL6R 358Ala allele impairs classical IL-6 receptor signaling and influences risk of diverse inflammatory diseases. PLoS Genet 9, e1003444.

54. Davis, W.A., Knuiman, M., Kendall, P., Grange, V., Davis, T.M.E., and Fremantle Diabetes Study (2004). Glycemic exposure is associated with reduced pulmonary function in type 2 diabetes: the Fremantle Diabetes Study. Diabetes Care 27, 752–757.

55. King, J.B., West, M.B., Cook, P.F., and Hanigan, M.H. (2009). A novel, species-specific class of uncompetitive inhibitors of gamma-glutamyl transpeptidase. J Biol Chem 284, 9059– 9065.

56. Tuzova, M., Jean, J.-C., Hughey, R.P., Brown, L.A.S., Cruikshank, W.W., Hiratake, J., and Joyce-Brady, M. (2014). Inhibiting lung lining fluid glutathione metabolism with GGsTop as a novel treatment for asthma. Front Pharmacol 5, 179.

57. Smailhodzic, D., van Asten, F., Blom, A.M., Mohlin, F.C., den Hollander, A.I., van de Ven, J.P.H., van Huet, R.A.C., Groenewoud, J.M.M., Tian, Y., Berendschot, T.T.J.M., et al. (2014). Zinc Supplementation Inhibits Complement Activation in Age-Related Macular Degeneration. PLoS ONE 9, e112682.

58. Nan, R., Tetchner, S., Rodriguez, E., Pao, P.-J., Gor, J., Lengyel, I., and Perkins, S.J. (2013). Zinc-induced self-association of complement C3b and Factor H: implications for inflammation and age-related macular degeneration. J Biol Chem 288, 19197–19210.

59. Bustamante-Marin, X.M., and Ostrowski, L.E. (2017). Cilia and Mucociliary Clearance. Cold Spring Harb Perspect Biol 9,.

60. Hewson, C.A., Haas, J.J., Bartlett, N.W., Message, S.D., Laza-Stanca, V., Kebadze, T., Caramori, G., Zhu, J., Edbrooke, M.R., Stanciu, L.A., et al. (2010). Rhinovirus induces MUC5AC in a human infection model and in vitro via NF-κB and EGFR pathways. Eur Respir J 36, 1425–1435.

61. Barbier, D., Garcia-Verdugo, I., Pothlichet, J., Khazen, R., Descamps, D., Rousseau, K., Thornton, D., Si-Tahar, M., Touqui, L., Chignard, M., et al. (2012). Influenza A induces the major secreted airway mucin MUC5AC in a protease-EGFR-extracellular regulated kinase-Sp1-dependent pathway. Am J Respir Cell Mol Biol 47, 149–157.

62. Singanayagam, A., Footitt, J., Kasdorf, B.T., Marczynski, M., Cross, M.T., Finney, L.J., Trujillo Torralbo, M.-B., Calderazzo, M., Zhu, J., Aniscenko, J., et al. (2019). MUC5AC drives COPD exacerbation severity through amplification of virus-induced airway inflammation (Immunology).

63. Lu, W., Liu, X., Wang, T., Liu, F., Zhu, A., Lin, Y., Luo, J., Ye, F., He, J., Zhao, J., et al. (2020). Elevated MUC1 and MUC5AC mucin protein levels in airway mucus of critical ill COVID-19 patients. J Med Virol.

64. He, J., Cai, S., Feng, H., Cai, B., Lin, L., Mai, Y., Fan, Y., Zhu, A., Huang, H., Shi, J., et al. (2020). Single-cell analysis reveals bronchoalveolar epithelial dysfunction in COVID-19 patients. Protein Cell 11, 680–687.

65. (2009). Essentials of Glycobiology (Cold Spring Harbor (NY): Cold Spring Harbor Laboratory Press).

66. Wang, S.-S., del Solar, V., Yu, X., Antonopoulos, A., Friedman, A.E., Agarwal, K., Garg, M., Ahmed, S.M., Addya, A., Nasirikenari, M., et al. (2020). Efficient Inhibition of O-glycan biosynthesis using the hexosamine analog Ac _5_GalNTGc (Biochemistry).

67. Bae, S.S., Chang, L.C., Merkin, S.S., Elashoff, D., Ishigami, J., Matsushita, K., and Charles-Schoeman, C. (2020). Major Lipids and Future Risk of Pneumonia: 20-Year Observation of the Atherosclerosis Risk in Communities (ARIC) Study Cohort. The American Journal of Medicine S0002934320306987.

68. Chien, Y.-F., Chen, C.-Y., Hsu, C.-L., Chen, K.-Y., and Yu, C.-J. (2015). Decreased serum level of lipoprotein cholesterol is a poor prognostic factor for patients with severe community-acquired pneumonia that required intensive care unit admission. J Crit Care 30, 506–510.

69. Antcliffe, D., Jiménez, B., Veselkov, K., Holmes, E., and Gordon, A.C. (2017). Metabolic Profiling in Patients with Pneumonia on Intensive Care. EBioMedicine 18, 244– 253.

70. Nelson, M.R., Tipney, H., Painter, J.L., Shen, J., Nicoletti, P., Shen, Y., Floratos, A., Sham, P.C., Li, M.J., Wang, J., et al. (2015). The support of human genetic evidence for approved drug indications. Nat Genet 47, 856–860.

71. Asimit, J.L., Hatzikotoulas, K., McCarthy, M., Morris, A.P., and Zeggini, E. (2016). Trans-ethnic study design approaches for fine-mapping. Eur J Hum Genet 24, 1330–1336.

