## Supplementary Methods and Results for "Genome-wide meta-analysis of pneumonia suggests a role for mucin biology and provides novel drug repurposing opportunities"

#### SUPPLEMENTARY RESULTS

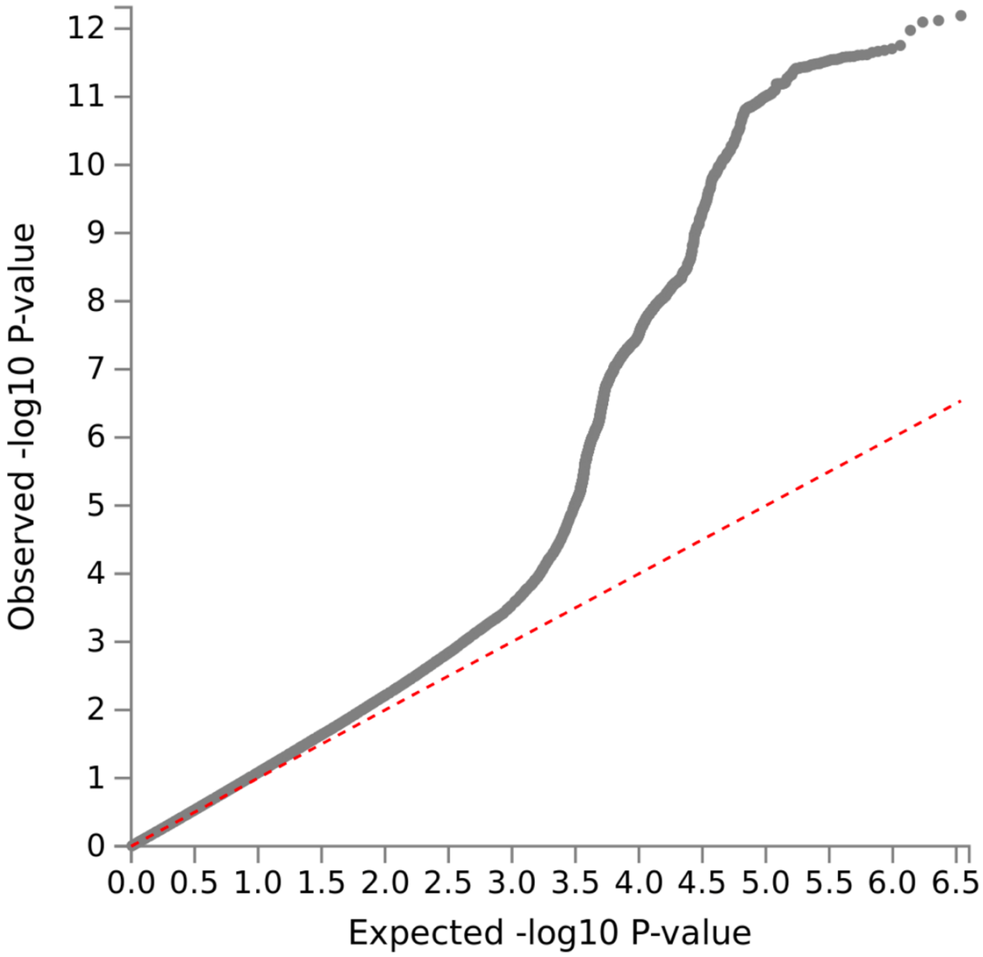


**Supplementary Figure 1. QQ-plot of common variant GWAS.** Quantile-quantile plot which visualises expected versus observed association, deviations from the diagonal line can be considered evidence of test-statistic inflation.

**Expanded description of novel common variant genome-wide significant loci associated with pneumonia susceptibility**

chr11:1110395-1232702 locus – lead SNP: rs28624253

This signal is located on chromosome 11 and is physiologically interesting given it is mapped to a region with a cluster of genes encoding mucin proteins (Supplementary Figure 2). Mucins are heavily glycosylated proteins that are produced in the epithelium and play an integral role in processes relevant to pneumonia, such as mucosal barriers to infection. The lead SNP rs28624253 is physically located upstream of one of the mucin genes in this region, *MUC5AC*. There were three other SNPs in this locus that were defined as independently significant SNPs using a very liberal criteria of $r^2 < 0.6$ with the lead SNP. Interestingly, these SNPs also had *MUC5AC* as their closest gene. Further support to this gene was provided by the fine mapping we performed through approximating ABFs (single causal variant assumption and prior variance of 0.2), wherein the 95% credible set spanned variants upstream or within *MUC5AC*. The highest CADD score (combined annotation dependent depletion) in this region, which is an integrated annotation method to assigning deleteriousness to variants, was rs1132436 (CADD score = 15.88), upstream of *MUC5AC* and annotated by Ensembl as a 'regulatory region variant'. In addition, the RegulomeDB framework scored this variant '2b' – which is indicative of some regulatory capacity. We also annotated variants in this locus based on their putative status as eQTLs from a collection of datasets curated by FUMA. There were seven genes implicated as a result (*MUC6*, *MUC5AC*, *MUC5B*, *CD151*, *EPS8L2*, *IFTM3*, and *RPLP2*), although it remains challenging to disentangle true eQTL effects from associations arising due to LD and further functional dissection is warranted to better interpret this locus. It should be noted that the eQTL signal for *MUC5AC* suggested that overexpression of this gene may increase the odds of pneumonia, that is, the pneumonia susceptibility risk allele is aligned with the alleles that are correlated with increased *MUC5AC* expression. However, this eQTL signal is derived from a post-mortem brain study of the dorsolateral prefrontal cortex from the PsychENCODE consortium, which is not likely a disease relevant tissue, although this sample is somewhat larger than many other eQTL datasets, and therefore, may have better power. Formal colocalisation analyses could be carried out in future work to assess whether the signal correlated with increased *MUC5AC* expression and increased odds of pneumonia are driven by the same causal variant/s. There was no evidence of long-range chromatin interactions encompassing this locus that would suggest the variants contained within exerted a distal effect.

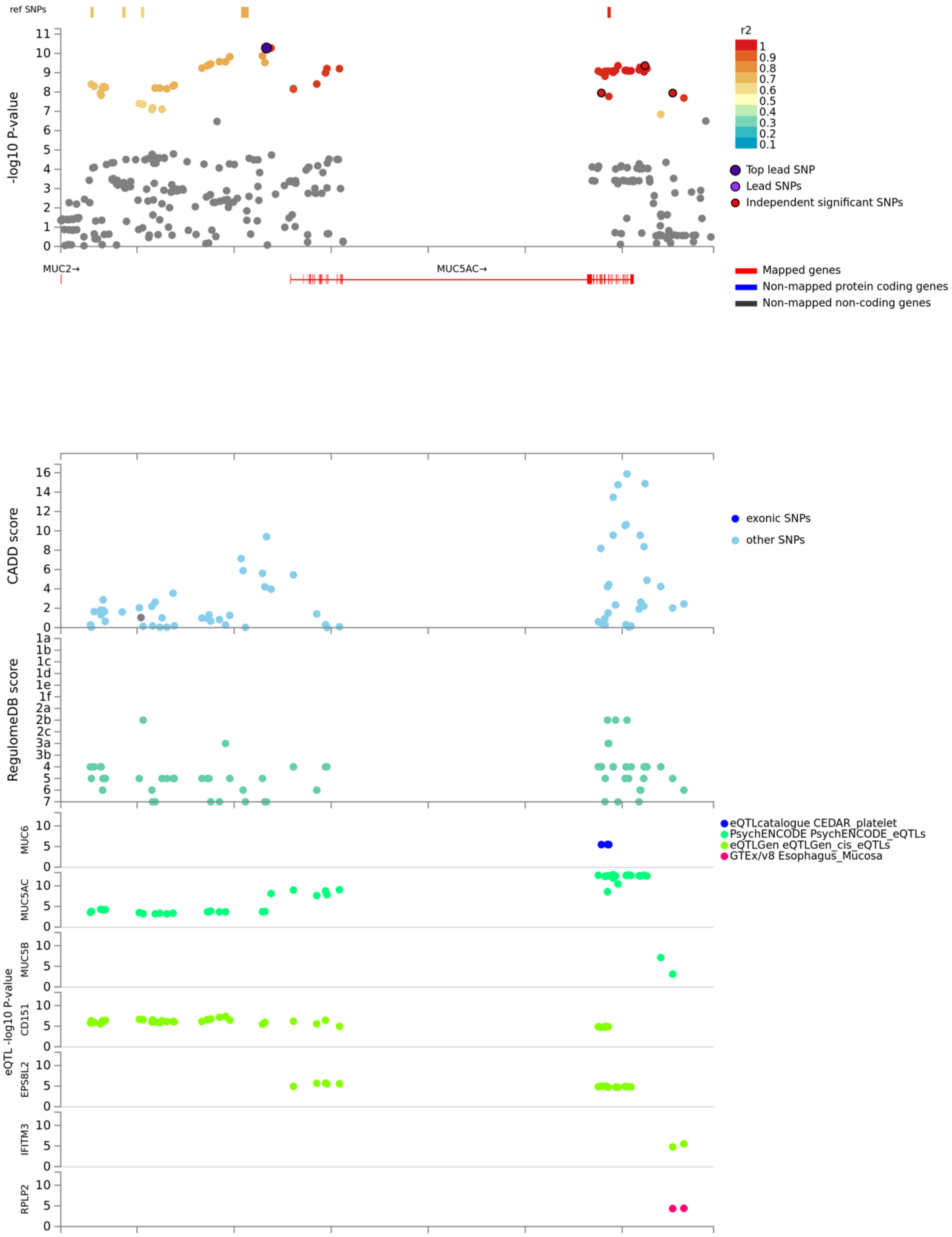


**Supplementary Figure 2. Region plot and variant annotation for the genome-wide significant pneumonia locus on chromosome 11.** The top panel plots the association of variants in this region as a negative log base 10 transformation of their *P* value. LD based on the UKBB release 2b 10k White British panel is used to colour the SNPs. The lead SNP is coloured purple, whilst independent significant SNPs ($r^2 <$ 0.6) are also highlighted. The next three panels are as follows, CADD score, RegulomeDB score, and eQTL association, with the tissue and consortia indicated by the legend.

chr10:76815686-76993015 locus – lead SNP: rs28624253

The locus of this second most significant novel pneumonia association beyond the MHC was physically mapped to a gene-rich region on chromosome 10 (Supplementary Figure 3a). The lead SNP itself was intergenic, with *PPIAP13* annotated as its closest gene. Moreover, the lead SNP displayed the highest posterior probability in the 95% credible set for this locus (*PP* = 0.676), whilst other variants in the credible set spanned genes such as *DUSP13* and *SAMD8*. Integrating Hi-C and eQTL data uncovered evidence of several genes that could be impacted by trait associated variation in this locus. Specifically, there were 11 genes for which expression was correlated with a variant in this locus (*C10orf11*, *ZNF503*, *COMTD1*, *VDAC2*, *SAMD8*, *DUSP13, DUPD1*, *KAT6B*, *ADK*, *AP3M1*, and *VCL*), whilst five of these also had evidence of chromatin interaction via Hi-C using lung, human embryonic stem cells, or mesenchymal stem cells – *COMTD1*, *VDAC2*, *KAT6B*, and *AP3M1* (Supplementary Figure 3b). As a result, this locus remains difficult to interpret in terms of pinpointing likely causal genes because of its complexity, however, the relevance of *VDAC2* was supported in this study by the TWAS and subsequent probabilistic finemapping of that signal. It is important to note that genes were only subjected to TWAS if they have a statistically significant model of genetically regulated expression, and thus, causal variation could be missed by only considering TWAS as a technique to pinpoint candidate genes from GWAS signals.

**a**

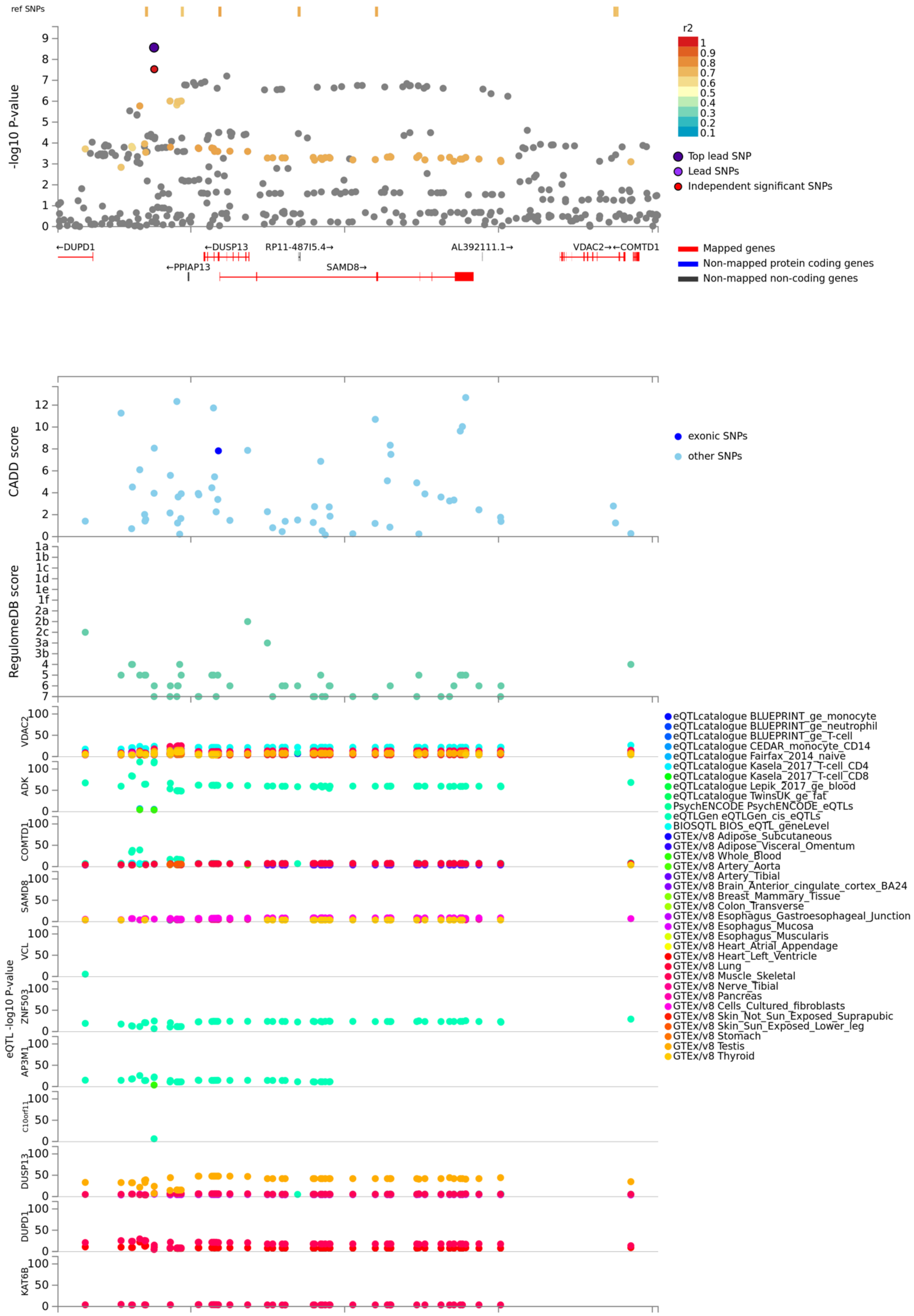

ref SNPs
-log10 P-value
r2
1
0.9
0.8
0.7
0.6
0.5
0.4
0.3
0.2
0.1
Top lead SNP
Lead SNPs
Independent significant SNPs
←DUPD1
←DUSP13
RP11-487I5.4→
AL392111.1→
VDAC2→←COMTD1
←PPIAP13
SAMD8→
Mapped genes
Non-mapped protein coding genes
Non-mapped non-coding genes
CADD score
exonic SNPs
other SNPs
RegulomeDB score
eQTL -log10 P-value
VDAC2
ADK
COMTD1
SAMD8
VCL
ZNF503
AP3M1
C10orf11
DUSP13
DUPD1
KAT6B
eQTLcatalogue BLUEPRINT_ge_monocyte
eQTLcatalogue BLUEPRINT_ge_neutrophil
eQTLcatalogue BLUEPRINT_ge_T-cell
eQTLcatalogue CEDAR_monocyte_CD14
eQTLcatalogue Fairfax_2014_naive
eQTLcatalogue Kasela_2017_T-cell_CD4
eQTLcatalogue Kasela_2017_T-cell_CD8
eQTLcatalogue Lepik_2017_ge_blood
eQTLcatalogue TwinsUK_ge_fat
PsychENCODE PsychENCODE_eQTLs
eQTLGen eQTLGen_cis_eQTLs
BIOSQTL BIOS_eQTL_geneLevel
GTEx/v8 Adipose_Subcutaneous
GTEx/v8 Adipose_Visceral_Omentum
GTEx/v8 Whole_Blood
GTEx/v8 Artery_Aorta
GTEx/v8 Artery_Tibial
GTEx/v8 Brain_Anterior_cingulate_cortex_BA24
GTEx/v8 Breast_Mammary_Tissue
GTEx/v8 Colon_Transverse
GTEx/v8 Esophagus_Gastroesophageal_Junction
GTEx/v8 Esophagus_Mucosa
GTEx/v8 Esophagus_Muscularis
GTEx/v8 Heart_Atrial_Appendage
GTEx/v8 Heart_Left_Ventricle
GTEx/v8 Lung
GTEx/v8 Muscle_Skeletal
GTEx/v8 Nerve_Tibial
GTEx/v8 Pancreas
GTEx/v8 Cells_Cultured_fibroblasts
GTEx/v8 Skin_Not_Sun_Exposed_Suprapubic
GTEx/v8 Skin_Sun_Exposed_Lower_leg
GTEx/v8 Stomach
GTEx/v8 Testis
GTEx/v8 Thyroid

**b**

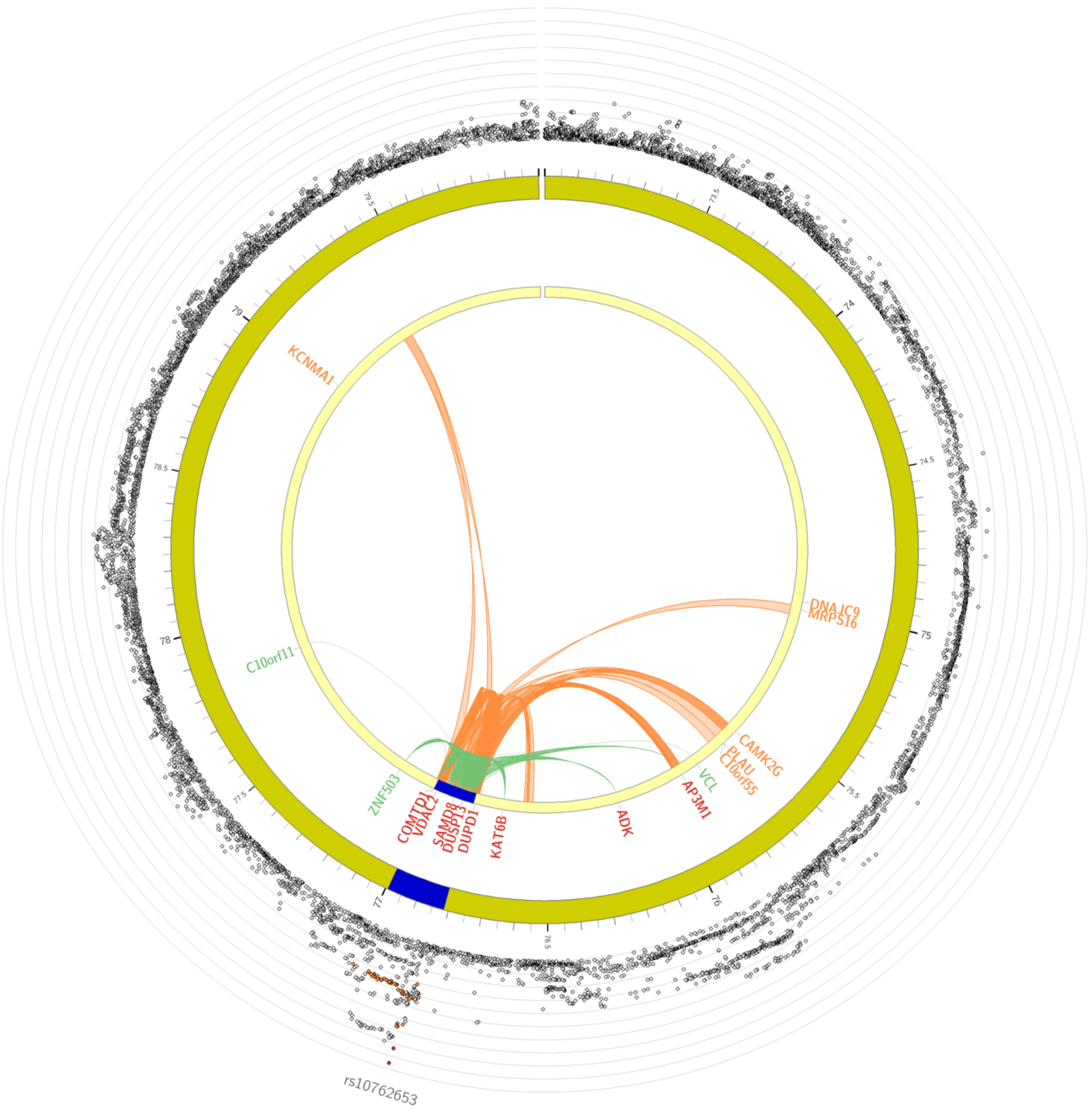


**Supplementary Figure 3. Region plot and variant annotation for the genome-wide significant pneumonia locus on chromosome 10.** (**a**) The top panel plots the association of variants in this region as a negative log base 10 transformation of their *P* value. LD based on the UKBB release 2b 10k White British panel is used to colour the SNPs. The lead SNP is coloured purple, whilst independent significant SNPs ($r^2 < 0.6$) are also highlighted. The next three panels are as follows, CADD score, RegulomeDB score, and eQTL association, with the tissue and consortia indicated by the legend. (**b**) Hi-C chromatin interactions on chromosome 10 for loci that reach genome-wide significance. The links are denoted by colour as follows: green = eQTLs in locus for that gene, orange = Hi-C interaction, red = eQTL and Hi-C.

<u>chr2:104056454-104380545 locus – lead SNP: rs264954</u>

The lead SNP for the third novel locus is located in an intergenic region, whilst the 95% credible set contains hundreds of variants, complicating the task of identifying candidate causal genes. However, several variants in the locus were annotated an eQTL in blood for *MFSD9*, a gene which is said to be a potential solute carrier. It should be noted that the association between variants in this region and *MFSD9* may be an artefact of linkage and further work is required to confirm this relationship.

**a**

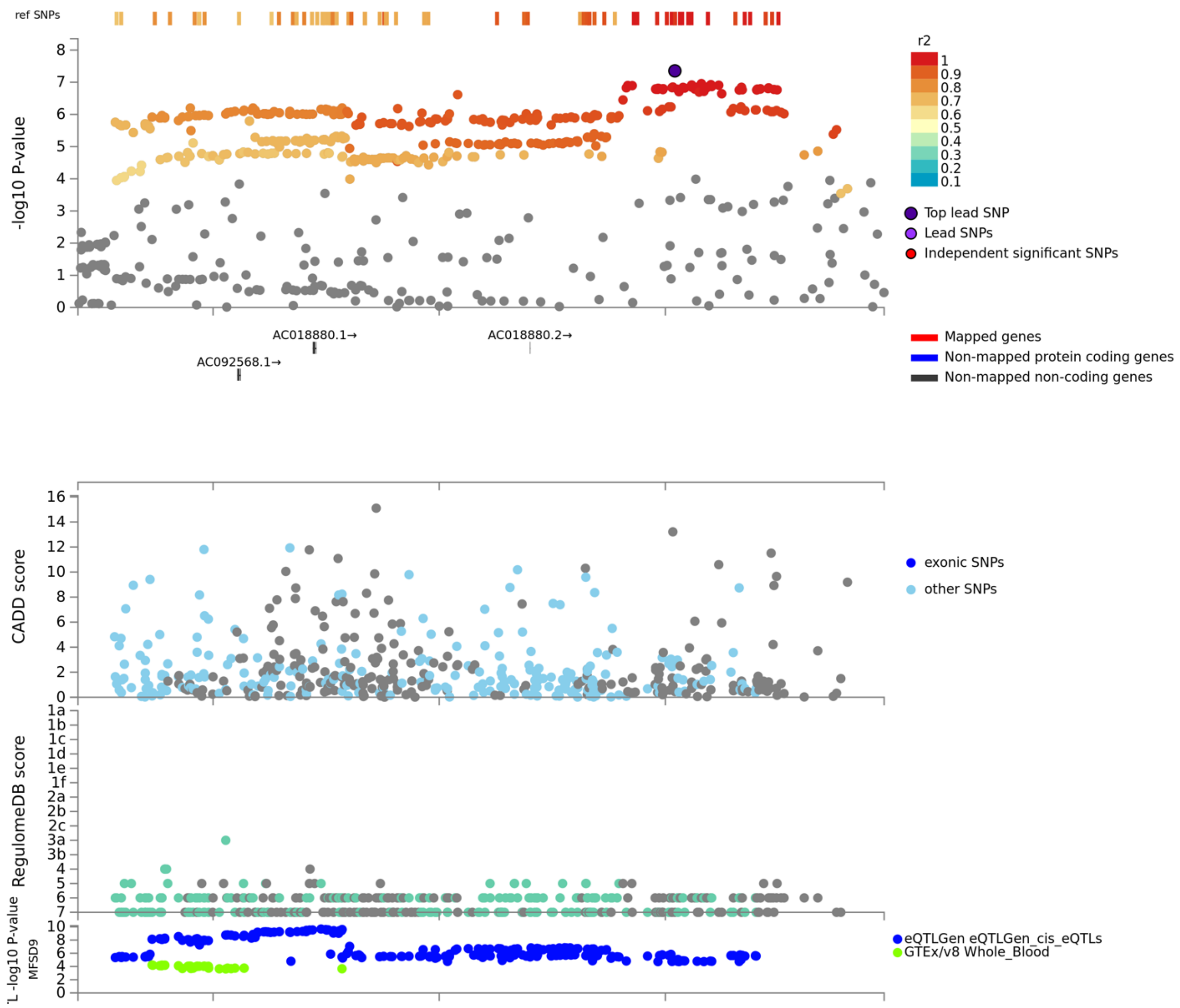

**b**

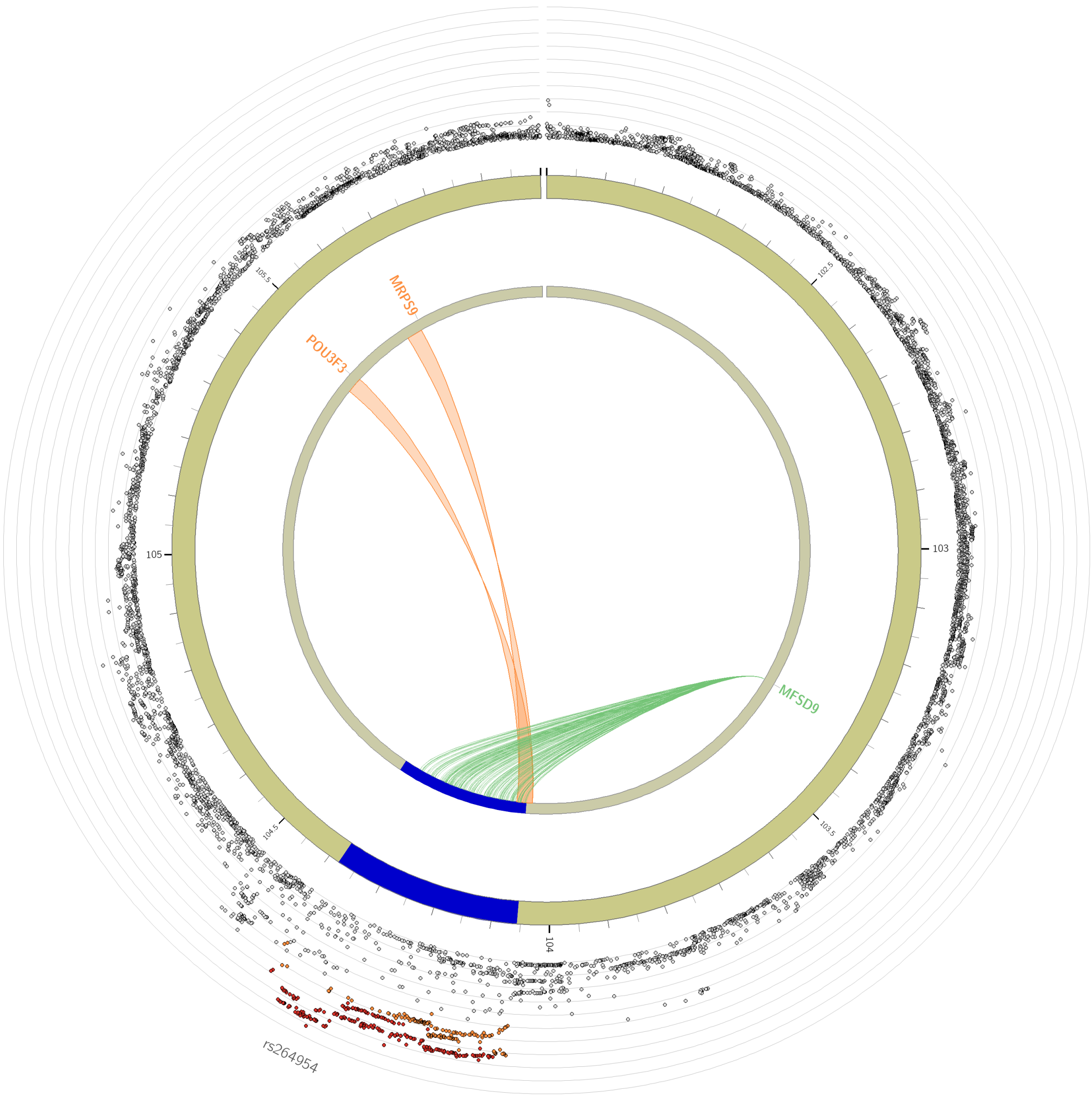


**Supplementary Figure 4. Region plot and variant annotation for the genome-wide significant pneumonia locus on chromosome 2.** The top panel plots the association of variants in this region as a negative log base 10 transformation of their *P* value. LD based on the UKBB release 2b 10k White British panel is used to colour the SNPs. The lead SNP is coloured purple, whilst independent significant SNPs ($r^2 < 0.6$) are also highlighted. The next three panels are as follows, CADD score, RegulomeDB score, and eQTL association, with the tissue and consortia indicated by the legend. (**b**) Hi-C chromatin interactions on chromosome 10 for loci that reach genome-wide significance. The links are denoted by colour as follows: green = eQTLs in locus for that gene, orange = Hi-C interaction, red = eQTL and Hi-C.

Gene prioritisation

We integrated a number of different annotation metrics in a tissue-specific and tissue agnostic manner to prioritise candidate genes from each novel genome-wide significant locus (Supplementary Figure 5). The criteria utilised in both configurations were: closest gene to the lead SNP, variant within 95% credible set mapped to gene, MAGMA gene-based association surviving Bonferroni correction, categorised as being a gene related to the pneumonia by DisGeNET (phenotype accession = C0032285), and an associated inflammatory or respiratory knock-out phenotype. Variants overlapping an eQTL signal and genes linked through chromatin interaction were considered for all tissues or for blood, lung, or spleen only.

Upon including eQTL and chromatin interaction information from any tissue, the highest confidence gene we found was *MUC5AC* given it satisfied the following: closest gene to the lead SNP, mapped to the 95% credible set of causal SNPs, overlapping an eQTL signal in the DLPFC (pneumonia odds increasing alleles overlaps with alleles correlated with increased *MUC5AC* expression), promoter anchored chromatin interaction between SNPs upstream of this gene and the *MUC5AC* promoter using DLPFC tissue, significant in the MAGMA gene-based analyses, linked to the pneumonia phenotype by DisGeNET, and has inflammatory and respiratory knock-out phenotypes. The eQTL and chromatin interaction was not seen using more disease relevant tissues, that is, lung, blood, or spleen, however, *MUC5AC* is relatively lowly expressed in most tissues, although, it is highly expressed in mucus-secreting goblet cells, and thus, further functional dissection of these specific cell types rather than bulk tissue may be required (1). Utilising only blood, lung, or spleen eQTLs and Hi-C data, *MUC5AC* remained the gene with the greatest number of lines of evidence, followed by *DUSP13*.

Genes with at least two lines of evidence in either the tissue-agnostic or tissue specific approach were subjected to pathway analysis using g:Profiler to identify gene-sets for which these genes display statistical overrepresentation after multiple testing correction (FDR < 0.05, Supplementary Figure 4C). We found a number of gene-sets that surpassed FDR < 0.05, including phenotypically relevant pathways such as *mucus layer*, *lung fibrosis*, and *O-linked glycosylation of mucins*.

**a**

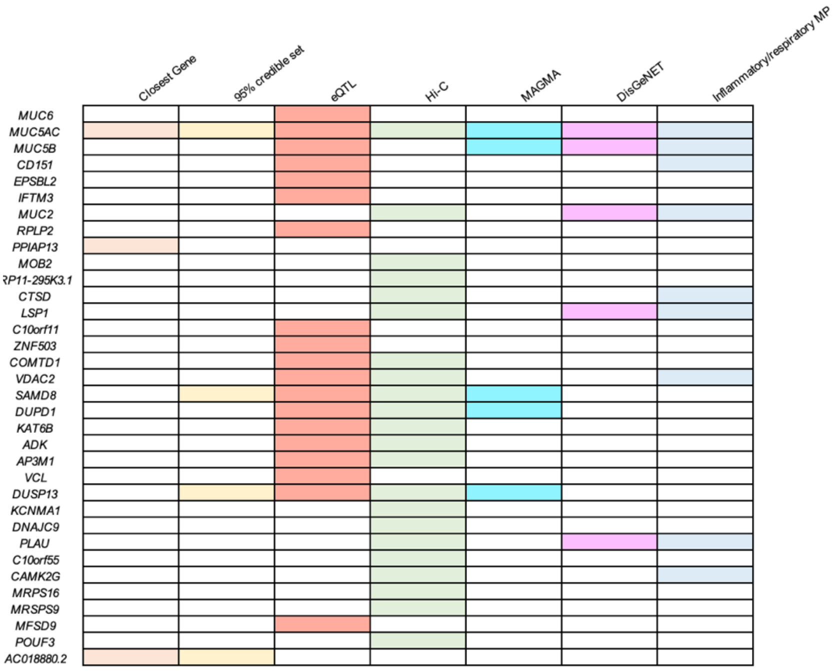


**b**

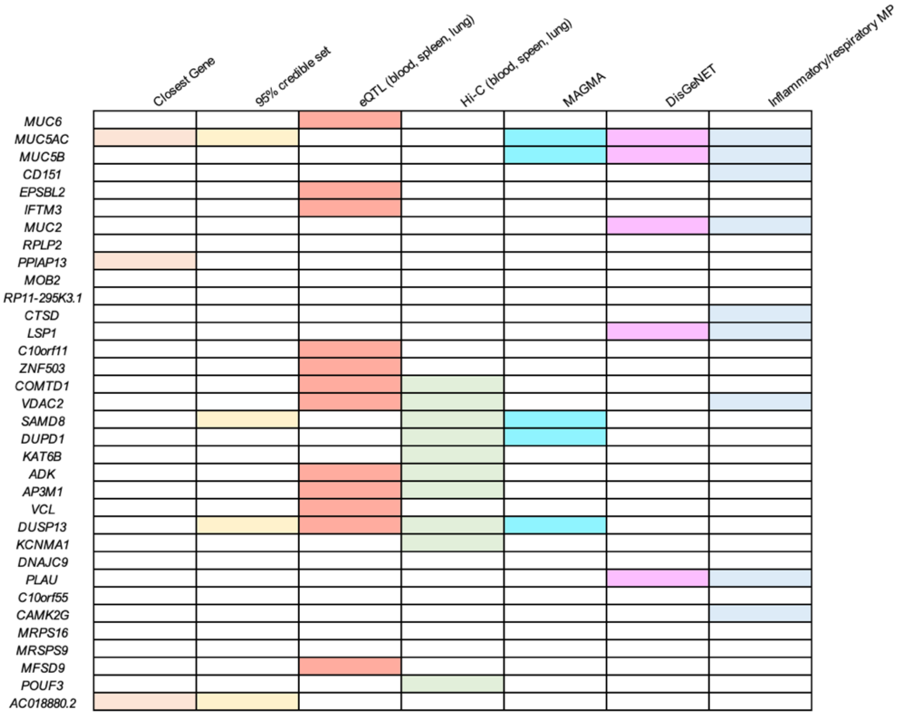


**c**

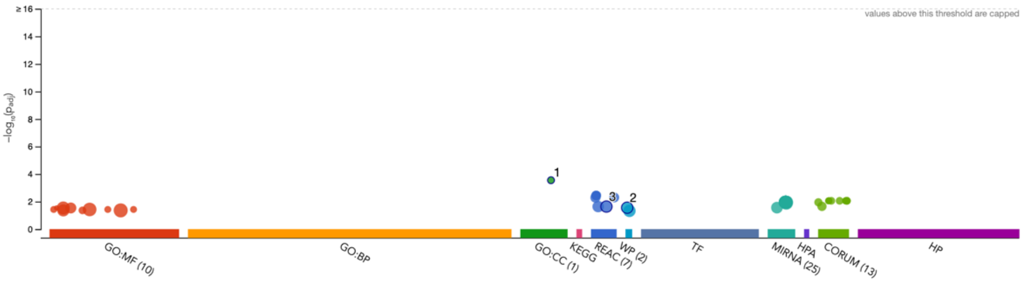


| ID | Source | Term ID | Term Name | $p_{adj}$ (query_1) |
|---|---|---|---|---|
| 1 | GO:CC | GO:0070701 | mucus layer | $2.804 \times 10^{-4}$ |
| 2 | WP | WP:WP3624 | Lung fibrosis | $2.771 \times 10^{-2}$ |
| 3 | REAC | REAC:R-HSA-9... | O-linked glycosylation of mucins | $2.294 \times 10^{-2}$ |

**Supplementary Figure 5. Gene prioritisation within genome-wide significant loci beyond the MHC region**. Each column in the table denotes an annotation metric, with each row a gene that satisfies at least one of the following criteria: closest gene to the lead SNP, gene mapped to SNPs in the derived 95% credible set, SNPs overlapping eQTL signal for gene, evidence of chromatin interaction (Hi-C) between SNP and gene, significant after correction in MAGMA gene-based analysis, associated with pneumonia phenotype by DisGeNET, evidence of an inflammatory or respiratory phenotype upon an *in vivo* knock-out. A shaded cell denotes that line of evidence. The eQTL and Hi-C data were obtained for (**a**) all tissues, and (**b**) lung, blood, and/or spleen only. (**c**) Gene overrepresentation analysis of genes with at least two lines of evidence from the above gene prioritisation, each point represents a gene-set that survives multiple-testing correction (FDR < 0.05) – the gene-set annotation categories were as follows: GO:MF = gene ontology, molecular function, GO:BP = gene ontology, biological process, GO:CC = gene ontology, cellular component, KEGG = KEGG pathways, REAC = REACTOME pathways, WP = WikiPathways, TF = transcription factor binding, MIRNA = microRNA with binding site, HPA = human protein atlas, CORUM = comprehensive resource of mammalian protein resources, and HP = human phenotype.

**Colocalisation of the mucin locus between adult-onset asthma and pneumonia susceptibility**

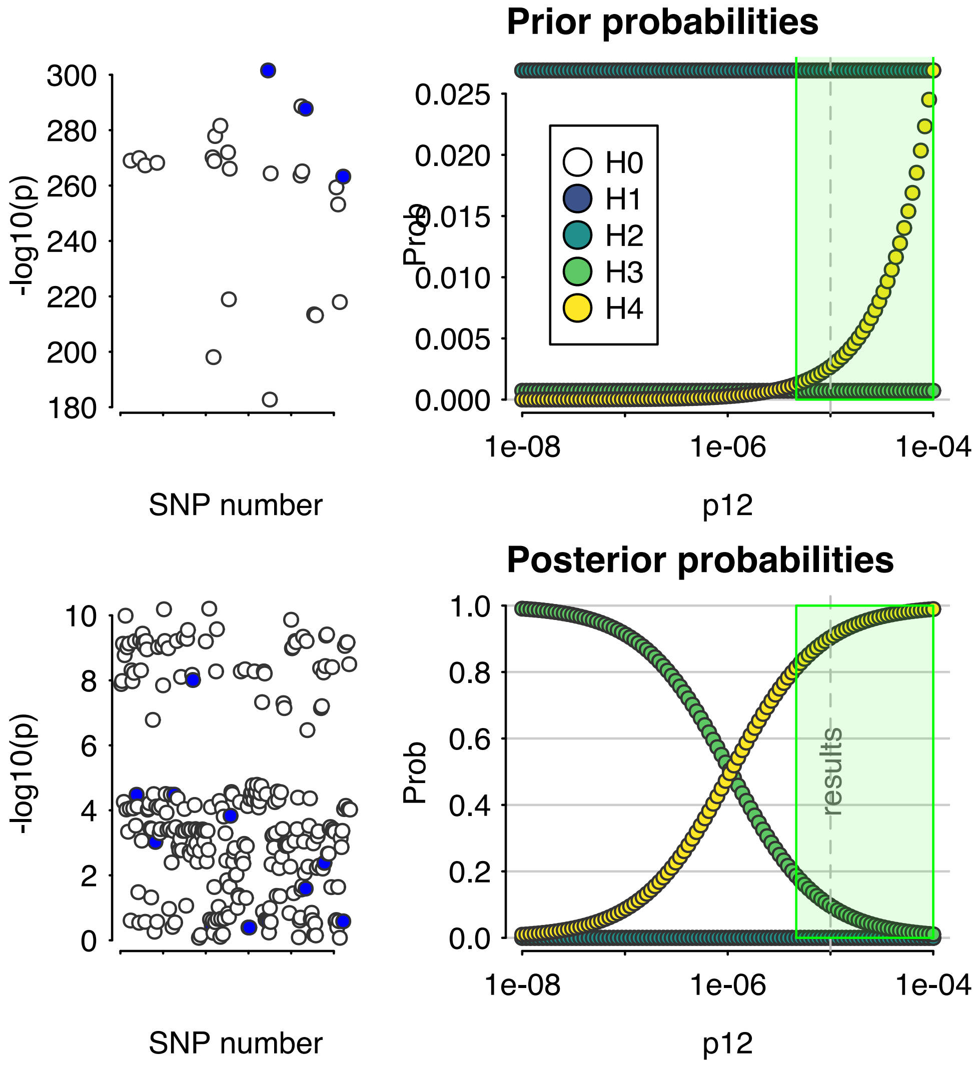


**Supplementary Figure 6. Sensitivity of the posterior probability of a shared causal variant for adult-onset asthma and pneumonia in the mucin locus to different priors.** We visualise the effect of using the default prior probability of hypothesis 4 ($H_4$), which was 1 x $10^5$ and demonstrate strong evidence for a shared causal variant – posterior probability > 90%. However, if a more conservative prior for $H_4$ was utilised, for example, < 1 x $10^{-6}$ lowers the posterior probability of $H_4$ and raises the posterior probability of $H_3$, which would indicate that the locus is associated with both traits but that there is a different underlying causal variant.

**Heterogeneity amongst SNP-pneumonia effect sizes between cohorts in the munged summary statistics**

As visualised below in supplementary figure 5, we found that considering the pneumonia summary statistics ‘munged’ to the HapMap3 panel, there were only a small number of variants that displayed suggestive association with pneumonia ($P < 1 \times 10^{-5}$) and significant heterogeneity.

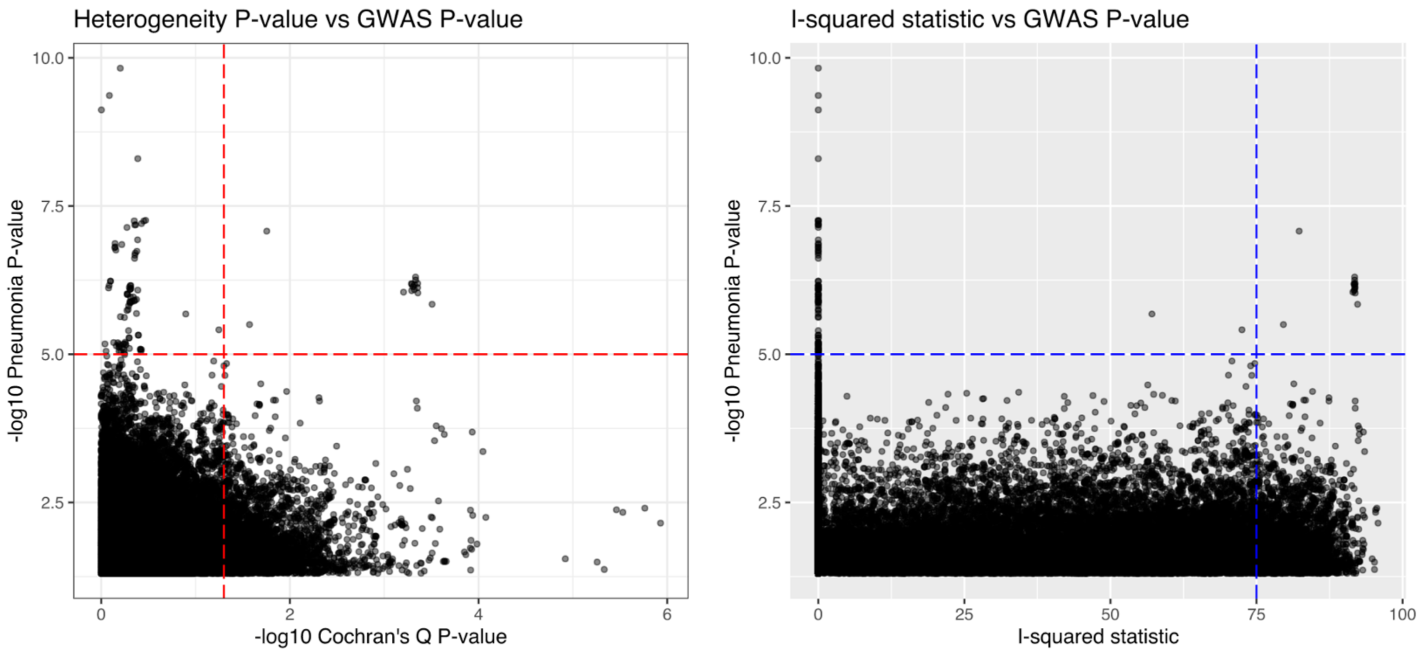


**Supplementary Figure 7. Genome-wide tests of heterogeneity of SNP pneumonia-effect sizes between the 23andMe and FinnGen cohorts.** We plot the two related metrics of heterogeneity (-$\log_{10}$ Cochran’s $Q$ $P$ value and $I^2$ statistic) -$\log_{10}$ SNP-pneumonia GWAS $P$ values for variants in the munged summary statistics (merged to the non-MHC HapMap3 panel) which displayed at least a nominal association with pneumonia ($P < 0.05$). We denote SNPs with suggestive significance ($P < 1 \times 10^{-5}$) on both plots with the horizontal dotted line and nominal evidence of heterogeneity (left plot: Cochran’s $Q$ $P < 0.05$, right plot: $I^2 > 75$).

### SUPPLEMENTARY METHODS

#### Study cohorts

The GWAS meta-analysis was performed using two primary study cohorts from 23andMe Inc. and FinnGen (release 3), respectively, as described below.

23andMe

Summary statistics for a self-reported pneumonia phenotype were obtained from 23andMe as outlined by Tian *et al.* (2). This self-reported phenotype was derived from an online survey of 23andMe customers about their medical history; specifically, the following two questions: "*Have you ever been diagnosed by a doctor with any of the following infectious conditions?" (Pneumonia: Yes, No, I don't know)*; *"Have you ever been diagnosed by a doctor with any of the following infectious conditions?"/"Pneumonia" (Never, 1-2 times, 3-5 times, More than 5 times, I'm not sure*). The endpoint for GWAS was binary such that individuals who answered yes to the first question, or at least once for the latter question, were cases, and those answering no or never, respectively, were controls. Moreover, cases could not have a negative response to one question and positive to the other, whilst controls were not permitted to have any positive responses. In the final GWAS after quality control (QC), there were 40600 cases and 90039 controls. The majority of self-reported pneumonia cases were female (56%), conversely, the majority of controls were male (54%). Participant age was described by four bins in the 23andMe meta-data for this GWAS: under 30, 30-45, 45-60, and over 60, with the oldest category the most common for cases (41%) and controls (31%). All individuals included in the 23andMe analyses provided informed consent and answered surveys online according to their human subjects protocol, which was reviewed and approved by Ethical & Independent Review Services, a private institutional review board (http://www.eandireview.com).

DNA was extracted from saliva samples and genotyped using one of four genotyping platforms, as described previously (2). This GWAS was restricted to unrelated individuals with European ancestry (2,3), with ancestry assigned by the *Ancestry composition* pipeline and relatedness tested through identity-by-descent (IBD) estimation (4). Imputation and phasing were performed separately for the four different genotyping platforms, with Beagle v3.3.1 utilised to phase the samples and Minimac2 leveraged for imputation to the March 2012 version three release of the 1000 genomes reference panel.

Genetic association in the 23andMe sample was performed using logistic regression covaried for age, sex, and the first four SNP derived principal components. This model assumed additive effects and *P* values were computed using a likelihood ratio test. Imputed dosages were utilised as opposed to 'best-guess' genotypes. Variants outside the pseudo-autosomal region on the X chromosome were coded as homozygous diploid for males. In our study, we retained only SNPs for meta-analysis that passed internal QC – specifically, SNPs were flagged and not retained if any of the following were met: *P* value for deviation from the Hardy-Weinberg equilibrium (HWE) of $P < 10^{-20}$, call rate < 90%, SNPs with a significant date of genotype effect (ANOVA of SNPs by a factor binning genotype data into 20 roughly equal bins), mitochondrial and Y chromosome SNPs, SNPs genotyped only on the first genotyping platform, an average imputation $R^2 < 0.5$ or a minimum $R^2 < 0.3$, and SNPs indicative of an imputation batch effect ($P < 10^{-50}$, ANOVA by a factor of imputation batch).

<u>FinnGen (release three)</u>

Summary statistics for pneumonia were downloaded from the third release of the FinnGen database which combines genotype data from Finnish biobanks and digital health record data from Finnish health registries. The pneumonia phenotype chosen was *All pneumoniae* (J10 pneumonia), for which 15771 cases and 119867 controls were available for GWAS after QC. This phenotype is level C in the ICD-hierarchy and encompasses several endpoint definitions which are as follows: viral pneumonia (Inflammation of the lung parenchyma that is caused by a viral infection) - Hospital Discharge registry/Cause of Death registry: ICD-10: J12, ICD-9: 480, ICD-8: 480; Pneumonia due to *Streptococcus pneumoniae* (streptococcal pneumonia: A febrile disease caused by streptococcus pneumoniae) - Hospital Discharge registry/Cause of Death registry: ICD-10: J13, ICD-9: 481, ICD-8: 48199; Pneumonia due to *Haemophilus influenzae* (bacterial pneumonia: Inflammation of the lung parenchyma that is caused by bacterial infections) - Hospital Discharge registry/Cause of Death registry: ICD-10: J14, ICD-8: 48210; Bacterial pneumonia, not elsewhere classified - Hospital Discharge registry/Cause of Death registry: ICD-10: J15, ICD-9: 482, ICD-8: 482; Pneumonia due to other infectious organisms, not elsewhere classified - Hospital Discharge registry/Cause of Death registry: I CD-10: J16, ICD-9: 483, ICD-8: 48399; Pneumonia, organism unspecified - Hospital Discharge registry/Cause of Death registry: ICD-10: J18, ICD-9: 485, ICD-8: 4850[2-9]|486. In the entire FinnGen cohort, the unadjusted prevalence of the *All pneumoniae* phenotype is

12.61%, with a mean age at first event of 53.81 and a case fatality rate at five years of 15.90%. The three drug classes most likely to be purchased after this diagnosis for the cohort were penicillins, fluroquinolones, and anilides.

The full genotyping, imputation, and GWAS protocol for release three of FinnGen has been outlined previously (https://finngen.gitbook.io/documentation/). Briefly, individuals from FinnGen contributing biobanks or cohorts were genotyped via Illumina or Affymetrix chip arrays. Individuals with sex mismatch, missigness > 5%, excess heterozygosity and non-Finish ancestry were removed. The final cohort consisted of unrelated, Finish ancestry individuals over the age of 18. Before imputation variants with call rate < 98%, a *P* value for deviation from HWE < $1x10^{-6}$, and minor allele count (MAC) < 3 were discarded, and pre-phasing performed using Eagle 2.3.5. Genotype imputation was undertaken using Beagle 4.1 and the population-specific SISu v3 reference panel that consists of 3775 high coverage whole genome-sequencing samples from six Finnish cohorts. The GWAS utilised the SAIGE model, which builds on the concept of logistic mixed models, covaried for age, sex, the first ten SNP-derived principal components, and genotyping batch (5). Association testing was performed for variants with a minimum MAC of 10.

**Meta-analysis**

The 23andMe and FinnGen summary statistics were meta-analysed using an inverse-variance weighted model with fixed effects as implemented by METAL version March 2011 (6). Firstly, we meta-analysed common variants, defined as sites with allele frequency > 1% in both the 23andMe and FinnGen cohorts. Variants were retained if they were available in both summary statistics and had an imputation quality that exceeded a minimum of 0.3 or a mean of 0.5 for variants not physically genotyped, resulting in 6888413 sites with an effect size estimate from the meta-analysis and a total sample size of 266277 individuals. Imputed rare variants available in both studies were subjected to a stricter filtering threshold for imputation quality such that only variants with a mean imputation quality > 0.5 or a mean value > 0.7 were subjected to meta-analysis, with 834366 low frequency variants considered. In both instances, we further tested for heterogeneity between the contributing studies using Cochran's *Q* test.

**Definition of a genome-wide significant locus**

Genome-wide summary statistics from the IVW meta-analysis were processed using the FUMA v1.3.6 (Functional Mapping and Annotation of Genome-Wide Association Studies) platform (7). Genome-wide significant variants were characterised using the traditional $P < 5 \times 10^{-8}$ threshold, whilst suggestive significance was defined a more lenient threshold of $P < 1 \times 10^{-5}$. We used the default settings for defining independent significant SNPs ($r^2 \leq 0.6$), followed by lead SNPs ($r^2 \leq 0.1$). The reference panel population for LD estimation was the UK biobank release 2b 10k White British panel, with LD blocks within 250 kb of each other merged into a single locus.

Genes within the genome-wide significant loci beyond the MHC region were then annotated to prioritise potential genes for further functional exploration. The following ranking criteria were implemented: gene closest to the lead SNP, genes mapped to SNPs that comprise part of the 95% credible set for each locus, SNPs overlapping eQTL signal for gene, SNPs overlapping locus of chromatin interaction with gene via Hi-C, gene significant using MAGMA, evidence of previous ontological and/or literature link to the pneumonia phenotype via DisGeNET v5.0 (8), and evidence of an inflammatory and/or respiratory phenotype in an *in vivo* knock-out model, leveraging the mouse genome informatics database (9). The eQTL and Hi-C studies included were curated by FUMA as described on their website.

**Rare variants associated with pneumonia at a suggestive significance threshold**

We further investigated variants with low frequency (Finish MAF < 0.01) that obtained suggestive significance in the pneumonia GWAS ($P < 1 \times 10^{-5}$). Variants were annotated using two online resources FAVOR (Functional Annotation of Variants; http://favor.genohub.org/) and VEP (Variant Effect Predictor; https://asia.ensembl.org/Tools/VEP).

**Estimation of SNP-based heritability**

SNP based heritability was computed using LD score regression (LDSR) with 1000 genomes phase 3 LD scores and weights (10). We converted the heritability estimate to the liability scale assuming the population prevalence of pneumonia of that of pneumonia in the FinnGen dataset (12.61%), as well as a more conservative population prevalence based on ICD-10 diagnosed pneumonia in the UK biobank (3.20% - see section: Pneumonia phenotype definition in the UK biobank cohort).

**Finemapping of genome-wide significant loci**

We finemapped the three-novel genome-wide significant loci outside of the MHC region by using a method which leverages approximated asymptotic Bayes' factors (ABF) to estimate credible sets under the assumption of a single causal variant (11,12). Specifically, we utilised Wakefield's method to approximate Bayes' factors assuming a prior variance of $0.2^2$, which reflects the belief that the confidence intervals of estimated variant effect sizes expressed as odds ratios range from around 0.68 to 1.48. Given that the posterior probability for causality of each variant is proportional to its Bayes' factor, these can be summed until a prespecified probability ($\rho$) is reached, thus constituting a '$\rho$ x 100%' set of putative causal variants.

**Impact of smoking and smoking heaviness on genetic associations with pneumonia**

We wished to investigate the potential confounding influence of genetic associations with smoking behaviour on the pneumonia genome-wide meta-analysis by conditioning the GWAS on two smoking related GWAS using the multi-trait-based conditional and joint analysis (mtCOJO) method (13). The smoking phenotypes selected were lifetime smoking initiation (ever vs never smoked, N = 262990) and smoking heaviness (cigarettes per day, N = 263954) (14). We recalculated heritability for the pneumonia summary statistics conditioned on lifetime smoking initiation and smoking heaviness separately, assuming the same population prevalence for liability scale conversion.

**Gene-based association**

Common variant (MAF > 0.01) SNP-wise *P* values were aggregated at gene-level using MAGMA v1.07b (15). Briefly, MAGMA combines *P* values using an adapted version of Brown's method whereby covariance between the SNPs is estimated based on LD from a population sample to scale the null $\chi^2$ distribution. Gene coordinates in hg19 assembly were obtained from NCBI and the 1000 genomes phase 3 European panel utilised as an LD reference. Genes within the MHC region were not considered due to the haplotype complexity of that region, as is usual practice. A window was set to extend the genic boundaries 35kb upstream and 10 kb downstream to capture potential regulatory variation. We compared this to a more conservative boundary definition of 5kb upstream and 1.5 kb downstream. The Bonferroni threshold for genic association was $P < 2.68 \times 10^{-6}$, accounting for the number of genes tested. Moreover, gene-based *P* values were leveraged for gene-set association using 1379 hallmark and canonical gene-sets from the Molecular Signatures Database (MSigDB) (16). After probit transformation of *P* to *Z*, a linear regression model was constructed such that

$Z$ was the outcome and binary indicator of gene-set membership an explanatory variable to test whether genes in the set were more associated than all other genes considered. This model was covaried for confounders including gene-size and MAC, as described elsewhere (15,17).

Rare variants (MAF < 0.01) were also aggregated at gene-level by leveraging the properties of the Cauchy distribution (18,19). Code for the Cauchy combination test was obtained from (https://github.com/yaowuliu/ACAT). The MAGMA approach for common variants accounts for dependency between $P$ values by estimating their covariance as a function of pairwise LD in a population sample – however, there are methodological challenges with this approach for rare variants and likely much larger samples would be required for accurate estimation, if any dependency between rare variants exists (20,21). Therefore, we employ an approach to combine $P$ values which guards against type I error inflation due to potential unknown covariance between rare variants. We acknowledge that this method may be conservative, particularly when $P$ values are larger (18). Rare variants were annotated to genes using ANNOVAR version 2017-07-17 (22). The test statistic ($T$) is a sum of $P$-values ($p_i$) transformed to approximate a Cauchy distribution, which is also flexible to incorporate weights ($w_i$; equation 1).

$$T = \sum_{i=1}^{k} w_i \tan\{(0.5 - p_i)\pi\} \quad (1)$$

Due to the heavy tail of the Cauchy distribution, $T$ is insensitive to correlations amongst the $P$ values, with the combined $P$ value approximated using the cumulative density function of the Cauchy distribution (equation 2). Two models were constructed: one using all genic variants and another including only those sites annotated as exonic. In both instances, genes required a minimum of two rare variants.

$$P_{Combined} \approx \frac{1}{2} - \left[\frac{\left\{\arctan\left(\frac{T}{w}\right)\right\}}{\pi}\right] \quad (2)$$

In addition, we constructed a model for rare variant gene-set association analogous to the MAGMA approach for common variants that leverages gene-based $Z$ scores (probit transformation of $P$; equation three). The same collection of pathways from MSigDB were considered, with genic $Z$ values regressed against a binary indicator of set membership ($\beta_S$), covaried for logarithmically transformed gene-length ($\beta_L$) and allele count ($\beta_C$). A one-sided test was performed for $\beta_S$, such that the null hypothesis is $\beta_S = 0$ and the alternative $\beta_S > 0$.

$$Z \sim \beta_0 + S\beta_S + L\beta_L + C\beta_C + \varepsilon \quad (3)$$

**Transcriptome-wide association studies of pneumonia**

A transcriptome-wide association study (TWAS) of pneumonia was performed using the FUSION method (23). TWAS tests the association between genetic variants comprising the models of predicted gene expression and the phenotype of interest. SNP weights were derived for genes with a significant contribution of *cis* acting SNPs to expression variability (*cis*-$h^2$ $P < 0.01$) using lung, whole blood, and spleen RNAseq GTEx v7 data. A transcriptome-wide significant gene was defined by the number of genes for which a TWAS $Z$ could be derived outside the MHC region, which was excluded, as follows: lung - $P < 7.05 \times 10^{-6}$ [$\alpha = 0.05/7095$], whole blood - $P < 9.24 \times 10^{-6}$ [$\alpha = 0.05/5414$], and spleen - $P < 1.22 \times 10^{-5}$ [$\alpha = 0.05/4107$]. The suggestive significance threshold was set as two-orders of magnitude higher than these $P$ values. For significant genes, we also assessed evidence for statistical colocalisation between the expression signal from the TWAS SNP weights and the association with pneumonia via the *coloc* methodology using default priors implemented in the FUSION.assoc_test.R script (24). Briefly, this method implemented the Bayesian finemapping approach described above to approximate ABFs for both the expression and GWAS association statistics for the SNP weights, respectively. The posterior probability (*PP*) of five hypotheses were calculated using this framework, specifically: $H_0$: no association with expression or pneumonia, $H_1$: association with expression but not pneumonia, $H_2$: association with pneumonia but not expression, $H_3$: association with expression and pneumonia, but two independent causal SNPs, and $H_4$: a single shared causal variant underlying the association between expression and pneumonia.

Another Bayesian method FOCUS was then utilised to finemap the TWAS associations which could be therapeutically useful (tier one or two) (25). Given observed TWAS statistics, the marginal posterior inclusion probability ($PIP$) was calculated and subsequently used to compute a credible set with 90% probability ($\rho$) of containing the causal gene ($c_i = 1$). As FOCUS allows the null model to be predicted as a possible member of the credible set, we excluded any genes for which that occurred. The credible set ($S$) was defined by summing normalised $PIP$ such that $\rho$ was exceeded, sorting the genes and then including those genes until at least $\rho$ of the normalized-posterior mass is explained (equation four).

$$S\,\{Gene_1, \dots, Gene_k\} = \sum_{i=1}^{k} PIP\ (c_i = 1|Z_{TWAS})\ \geq\ \rho \quad (4)$$

The Bernoulli prior for each causal indicator was set as the default $p$ = 1 x $10^{-3}$, with a default prior variance for effects at causal genes set as 40 ($n\sigma_c^2$ = 40). Previous work has demonstrated that FOCUS computed $PIP$s were robust to different specified prior variances (25). In all instances, we utilised a multi-tissue panel obtained from FOCUS GitHub repository which combines GTEx v7 SNP-weights with other FUSION TWAS weights (https://github.com/bogdanlab/focus/wiki, GTEx v7 with METSIM, CMC, YFS, and NTR).

**Genetic correlation and causal inference**

Bivariate linkage disequilibrium score regression (LDSR) was performed between pneumonia and a variety of GWAS as implemented by LDhub v1.9.3 (26). Summary statistics from the pneumonia meta-analysis were cleaned ('munged') prior to LDSR using munge_sumstats.py and merged with common HapMap3 SNPs excluding the major histocompatibility complex (MHC) region due to its LD complexity, as is usual practice (10). We retained estimates of genetic correlation ($r_g$) for GWAS with European ancestry and a heritability $z$ value > 4, as calculated by LDhub.

Latent causal variable models (LCV) were constructed between the most significantly correlated trait with pneumonia from each phenotypic category after multiple-testing. The RunLCV.R and MomentFunctions.R scripts were leveraged to perform these analyses (https://github.com/lukejoconnor/LCV). The LCV framework assumes that a latent variable, $L$, mediates the genetic correlation between two traits (trait one, trait two), and uses the mixed fourth moments of the bivariate effect size distribution to estimate the mean posterior genetic causality proportion (GCP) as described in detail by O'Connor and Price (27). The GCP estimate quantifies the magnitude of genetic causality between the two traits. GCP values range from −1 to 1 (full genetic causality), within these limits positive values indicate greater partial genetic causality of trait one on two, and vice versa for negative values. All traits were munged prior to LCV analyses, with only HapMap3 SNPs (MAF > 0.05) outside the MHC region retained in accordance with the LDSR analyses. We utilised the baseline 1000 genomes phase 3 LD scores for HapMap3 SNPs (MHC excluded). A two-sided $t$ test was used to assess whether the estimated GCP was significantly different from zero, with an absolute posterior mean GCP > 0.6 considered a reliable estimate.

We further interrogated the evidence of partial genetic causality between HDL and pneumonia by constructing a multivariable mendelian randomisation (MVMR) model leveraging instrumental variables for HDL, LDL, and triglycerides as implemented in the TwoSampleMR package version 0.5.5 (28). This model utilised IVs for the three lipid classes from Willer *et al.* (29) and calculates an exposure-outcome estimate for each lipid class conditioned on the remaining two.

**Drug repurposing**

There were three main strategies utilised for drug repurposing in this study: 1) mapping approved drug-gene interactions to drugs physically located within loci that surpassed suggestive significance for association with pneumonia ($P < 1 \times 10^{-5}$), 2) identification of biochemical factors through genetic correlation and causal inference that could be targeted by approved drugs in pneumonia, and 3) the *pharmagenic enrichment score* (PES) method for precision drug repurposing. Each of these three methods will be discussed in detail below:

<u>Mapping genes within suggestive pneumonia loci ($P < 1 \times 10^{-5}$) to known drug targets</u>

We identified all genes within suggestively significant loci as defined by FUMA outside the MHC, the MHC region was excluded to the large number of genes in this locus and the difficulty is disentangling the pneumonia signal for extensive linkage that the MHC displays. These non-MHC genes were then searched for evidence that they were targeted by an approved compound using the drug-gene interaction database (DGIdb) version 4.2.0, which curates drug-gene interaction data from multiple databases, such as DrugBank and ChemblInteractions, along with curated information from the literature (30). We retained approved compounds that had at least three lines of evidence for targeting one of our genes of interest.

<u>Biochemical traits as drug targets for pneumonia</u>

The biochemical GWAS interrogated to estimate genetic correlation and infer potential causal relationships were a collection of GWAS of blood based biochemical traits collected from UK biobank participants, including, lipids, hormones, blood cell counts, and vitamins.

These GWAS were performed by Ben Neale's group and were publicly available for download from the group's website (http://www.nealelab.is/uk-biobank). Specifically, we selected 50 of these traits for which the estimate of SNP heritability was significantly different from zero and categorised as 'high' or 'medium' confidence estimates. The full details of these categorisations have been outlined elsewhere (https://nealelab.github.io/UKBB_ldsc/confidence.html).

In all instances, we utilised the GWAS summary statistics for which the phenotype of interest was subject to inverse-rank normal transformation. The covariates for these GWAS were age, age$^2$, sex, age x sex, age$^2$ x sex, and the first 20 principal components. We used LDSR to estimate genetic correlation with each of these phenotypes, as above. Thereafter, LCV models were constructed between pneumonia and the biochemical traits that displayed significant genetic correlation after multiple testing correction.

MR was then implemented to further probe the relationships between triglycerides and pneumonia, and gamma glutamyl-transferase (GGT) and pneumonia. Firstly, we attempted to replicate the lipid MVMR model by utilising IVs for HDL, LDL, and triglycerides from the UK biobank rather than the Willer *et al.* GWAS. Univariable MR was then undertaken for the effect of triglycerides and GGT on pneumonia. The primary model was an inverse-variance weighted (IVW) estimator (31), with a comparison made between a model with fixed effects versus multiplicative random effects. The random effects approach is better suited to instances with IV exposure-outcome effect heterogeneity, as was tested with Cochran's *Q* test (32). The IVW model is considered to be the most well-powered out of the suite of MR methods we implemented in this study, however, it makes a potentially unrealistic assumption that all IVs are valid. The use of a genetic variant as an IV is underpinned by three central assumptions, as has been discussed extensively elsewhere (33,34):

IV1: the variant is rigorously associated with the exposure;

IV2: the variant is independent of all confounders of the exposure-outcome relationship ;and,

IV3: the variant is associated with the outcome only by acting through the exposure (independent conditional on the exposure and confounders).

It should be noted that only the first assumption can be directly tested, as a result, we utilised four additional MR methods as a sensitivity analysis that make different underlying assumptions about IV validity. Firstly, a weighted median estimator that is subject to the ‘majority valid’ assumption (unbiased estimate as long as < 50% of IVs are invalid) by taking the median of ratio estimates rather than the mean like the IVW method (33). Secondly, a mode-based estimator was implemented that is underpinned by the related ‘plurality valid’ assumption (35). Thirdly, an MR-Egger model was constructed which is an adaption of Egger regression wherein the exposure effect is regressed against the outcome with an intercept term added to represent the average pleiotropic effect (34). Finally, we utilised the outlier robust MR-Pleiotropy Residual Sum and Outlier (MR-PRESSO) framework (36). The advantage of this method is that it seeks to identify outlier IVs by considering the residual sum of squares (RSS, a heterogeneity measure of IV estimates) and testing for deviations from a simulated Gaussian distribution of expected RSS when each IV is iteratively removed. We note that no outliers were identified in the triglyceride or GGT model. Three additional statistical methods were implemented to test for evidence of potential confounding pleiotropy: testing whether the Egger intercept is significantly different from zero, the MR-PRESSO global pleiotropy test, and a leave-one-out analysis; whereby each IV was iteratively removed and the IVW estimate recalculated to evaluate evidence of disproportionately large IV effects that may be indicative of a pleiotropic signal. It is of critical importance that researchers interpreting the output of these statistical methods to evaluate pleiotropy are cognisant that it is not possible to exclude a confounding influence of horizontal pleiotropy through statistical methods alone.

Moreover, we investigated the observational association between triglyceride and GGT concentration amongst individuals from the UK biobank without missing values for the respective biochemical trait or covariate values. The definition of the pneumonia phenotype in the UK biobank is discussed in the subsequent section of this document (*PES analyses in the UK biobank*). The measures of triglycerides and GGT as baseline was scaled to have zero mean and unit variance. We constructed a binomial logistic regression model between scaled triglycerides, and subsequently GGT, and pneumonia, covaried for sex, age, age$^2$, sex*age, sex*age$^2$, smoking status (ever vs never smoked), and Townsend deprivation index. Furthermore, we also tested the association between individuals in the 90$^{th}$ percentile for each measure and pneumonia diagnosis relative to the remaining participants. The final sample sizes were 466,307 for the triglyceride models, and 466,435 for the GGT analyses. Furthermore, we repeated the test between triglyceride or GGT levels and pneumonia diagnosis amongst a subset of the UK biobank cohort who have relatively lower risk of pneumonia, specifically, females aged under 45 at the time of assessment who are self-reported lifetime non-smokers.

The sample sizes were 14,953 for the triglyceride cohort, and 14,947 for the GGT cohort, with the logistic regression model here still covaried for Townsend deprivation index.

#### The *pharmagenic enrichment score* (PES) framework

The *pharmagenic enrichment score* (PES) is an approach to perform genomics-informed precision drug repurposing (37). Specifically, pathways (gene-sets) with known drug targets are identified which display some evidence of harbouring an enrichment of trait associated variation relative to all other genes. These pathways then become the basis for a genetic risk score using only those variants mapped to genes within the pathway (PES). Individuals with high genetic risk within a particular druggable pathway (elevated PES) may benefit from a drug which modulates that pathway. We identified candidate gene-sets to construct pneumonia PES by using a modified version of MAGMA. We implemented *P* value thresholding in the calculation of the gene-based test statistic, as we have described previously (37,38). Briefly, the underlying concept of this is that there may be different biological insights that can be gained by focusing on pathways enriched with variations at different levels of the polygenic signal. For example, consider a pathway which displays enrichment when all variants are considered ($P < 1$), versus a pathway which only displays enrichment when nominally significant variants are included ($P < 0.05$). The nominally significant variants represent a less 'polygenic' signal relative to all variants, and thus, may encompass distinct biological processes. We posit that this concept is somewhat analogous to the *P* value thresholding when selecting the most parsimonious PRS configuration, whereby different *P* value thresholds seem to fit better depending on the target phenotype of interest. In accordance with our previous work we somewhat arbitrarily selected four *P* value thresholds which balance capturing different elements of the polygenic signal with encompassing enough SNPs to construct scores using variants only within a biological pathway, which may not be realistic at lower *P* thresholds – specifically, we chose, $P < 1$ (all SNPs), $P < 0.5$, $P < 0.05$, and $P < 0.005$ (37). The definition of a 'druggable' biological pathway has been described elsewhere (17), briefly the MSigDB gene-sets used for gene-set association in this study were processed to identify a subset of gene-sets (N=1030) with at least one high confidence gene targeted by an approved pharmacological agent.

We selected pathways that survived multiple-testing correction for an enrichment of pneumonia associated variation relative to all other genes at that threshold by applying correction via the Benjamini-Hochberg (BH) method (FDR < 0.05) to all thresholds combined. These associations can be interpreted based on the *P* value threshold for the model, for example, at gene-set which survives FDR correction that includes only variants which displayed a nominally significant univariable association with pneumonia ($P < 0.05$) is indicative of a set of genes that are more associated with pneumonia than all other genes with at least one SNP that had $P < 0.05$ in the GWAS. The BH approach was implemented rather than Bonferroni as several gene-sets will be tested multiple times at different *P*-value thresholds, and thus, the assumption of independence underlying Bonferroni correction likely means this would be overly conservative. These pathways that survived correction were subjected to two approaches to identify drug repurposing candidates by virtue of targeting a gene or genes in these pathways. We have outlined these approaches in detail elsewhere (37,38). Firstly, drugs with at least three lines of evidence of targeting one or more genes from each pathway were retained by leveraging data in the drug-gene interaction database (DGIdb v4.2.0) (30), along with DrugBank compounds that interact with more genes in any of the pathways than would be expected by chance after correcting for the number of compounds tested using the BH approach (FDR < 0.05). The latter analyses were performed using the WebGestalt R package v 4.0.2 (39).

PES analyses in the UK biobank cohort

*Pneumonia phenotype definition in the UK biobank cohort*

We utilised the UK biobank (UKBB) cohort to test the association between a pneumonia PRS and pneumonia phenotypes recorded for these participants. The UKBB is a large, longitudinal study that has recorded extensive phenotypic information accompanied by genetic data for the majority of individuals (40). Individuals for whom the following information was non-missing were retained (N = 498,989) – phenotypic sex (field 31.0.0), age at assessment (field 21003.0.0), self-reported smoking history (ever vs never smoked, field 20160), and the Townsend deprivation index metric (field 189.0.0).

There were two main criteria used to define our set of lifetime-pneumonia cases, with a strict and broad definition used. Firstly, we identified individuals who self-reported a pneumonia diagnosis at any of the assessment visits. Secondly, we leveraged primary and secondary ICD-

10 codes for each participant from hospital inpatient records. The ICD-10 codes used to define the pneumonia phenotype were as follows: J100 – influenza with pneumonia, influenza virus identified, J110 – influenza with pneumonia, virus not identified, J12 – viral pneumonia, not elsewhere classified, J13 - Pneumonia due to *Streptococcus pneumoniae*, J14 - Pneumonia due to *Haemophilus influenzae*, J15 - Bacterial pneumonia, not elsewhere classified, J16 - Pneumonia due to other infectious organisms, not elsewhere classified, J17 - Pneumonia in diseases classified elsewhere, and J18 - Pneumonia, organism unspecified. There were 15,725 individuals from the cohort with a primary or secondary diagnosis using the ICD-10 schema for at least one of the above codes. In the strict phenotype definition, we defined cases as those satisfying ICD-10 criteria as above, and controls as all those who did not have one of those codes recorded along with any individual who self-reported pneumonia without a pneumonia ICD-10 code ($N_{Controls}$ = 476,677). Individuals who self-reported pneumonia but were not hospitalised likely were diagnosed with the disease but we removed them in this strict configuration. In the broad-phenotype definition, pneumonia cases were individuals with a relevant ICD-10 code or a self-reported lifetime pneumonia diagnosis (N = 22,312).

*Genotyping, imputation, and quality control of the UK biobank SNP array data*

The genotyping and imputation procedures for UKBB participants have been described extensively elsewhere (41). We obtained chromosome-wise imputed data in Oxford bgen format from the UKBB as per our application (version 3 imputation) and restricted variants to sites in the Haplotype Reference Consortium panel (~40 million variants). Sample exclusions comprised of any individuals who satisfied one or more of the following criteria – missing sex recorded at baseline (field 31), mismatch between recorded sex and genetically inferred sex (field 22001), evidence of sex chromosome aneuploidy (field 22019), excess heterozygosity and missing rate (field 22027), ten or more third-degree relatives identified in the sample (field 22021), exclusion from the kinship inference process (field 22021), and other flagged sample exclusions in the UKBB meta-data (field 22010), . Thereafter, unrelated individuals were retained by virtue of being in the principal components analysis (PCA) conducted by the UKBB (as these individuals were deemed unrelated by the UKBB). The analyses in this manuscript were restricted to a homogeneous white-British subset of the UKBB to attempt to guard against unwanted effects of population stratification. This subset was ascertained by selecting those participants with self-identified 'white British' ancestry and a very similar genetic ancestry based on the projection of eigenvectors from the PCA in the work done by the UKBB (41).

Post-imputation QC was as follows and performed using PLINK 2 – firstly, well-imputed variants were retained using a threshold of variant INFO > 0.8, followed by excluding variants that satisfied or more of the following: MAF < $1x10^{-4}$, strong deviation from the Hardy-Weinberg equilibrium ($P < 1 \times 10^{-10}$) and call rate < 0.98. This resulted in a final set of 336,896 participants and 13,568,914 variants that survived all of the above QC. We reperformed PCA using this filtered white British ancestry subset using FlashPCA2 v2.0 (42) in order to calculate eigenvectors to include as covariates in downstream analyses. As is usual practice, we only included variants with MAF > 0.05 in relative linkage equilibrium (pairwise $r^2 < 0.05$), that were physically genotyped on both array types and were not in regions of long-range LD known to confound PCA, such as the MHC region on chromosome six (43).

*Generation of PES and PRS profiles*

We excluded the MHC region from the PES and genome wide PRS due to the complexity of this region, as is usual practice. Variants were mapped to each gene comprising the PES pathway if they were within the defined gene coordinates, or in the genic boundary used for the identification of the PES gene-sets, that is, 35 kb upstream or 15 kb downstream of the gene. PRSice v.2.3.3 was used to calculate the scores per autosome, which were then summed in R to obtain the full scores. The *P* value threshold for variant inclusion in the PES was the same as the threshold used to identify the pathway in the gene-set association. We utilised the default LD $r^2$ for PRSice-2 ($r^2 < 0.1$), with the PES calculated under an additive model whereby the effect size of each independent variant in the pathway was multiplied by its zygosity and summed for each individual. The same framework was applied as above to derive genome wide PRS using the following *P* value thresholds ($P < 1 \times 10^{-5}$, $P < 0.005$, $P < 0.05$, $P < 0.1$, $P < 0.5$, and $P < 1$).

*Association tests for PES and PRS*

There were 10,540 individuals from the genotyped subset of the cohort included in the PES calculation with a primary or secondary diagnosis using the ICD-10 primary or secondary diagnosis codes relevant to pneumonia. In the strict phenotype definition, we defined cases as those satisfying ICD-10 criteria. and controls as all those who did not have one of those codes recorded along with any individual who self-reported pneumonia without a pneumonia ICD-10 code ($N_{Controls}$ = 320,213). Individuals who self-reported pneumonia but were not hospitalised likely were diagnosed with the disease but we removed them in this strict configuration. In the broad-phenotype definition, pneumonia cases were individuals with a relevant ICD-10 code or a self-reported lifetime pneumonia diagnosis (N = 15,138). The association between each PES and PRS (scaled to zero mean and unit variance) and either of these pneumonia phenotypes was tested using binomial logistic regression adjusted for age, age$^2$, sex, genotyping batch, and the first ten SNP derived principal components. The PRS $P$ value threshold with the most variance explained was selected using Nagelkerke's $R^2$. Given the putative relationship between triglycerides and bile acid metabolism, we also tested the association between the *Bile acid metabolism* PES and PRS with a linear model using the same covariates. A sensitivity model was also constructed with the addition of statin use as a covariate. Briefly, we defined statin use as a binary variable in the UKBB using the self-reported medication data (field 20003), and defined statin use as individuals who self-reported any of the following – atorvastatin usage, fluvastatin usage, pravastatin usage, rosuvastatin usage, and/or simvastatin usage. The triglyceride values were scaled to have zero mean and unit variance, as done previously in the manuscript. In addition, we constructed two other models to test the sensitivity of the *Bile acid metabolism* PES ~ triglyceride concentration correlation, a model with an additional covariate of PRS at the same $P$ value threshold as the PES, and a triglyceride outcome variable that was winsorized at three standard deviations above the cohort mean before normalisation.

We obtained multiplexed IgG antibody data for a random subset of the UKBB for whom this pilot study was performed, of this subset, 6,443 participants were in our white British ancestry genotyped cohort. Full details of the platform on which these antibody data were processed has been described elsewhere (44). We selected 14 pathogens where seropositivity > 5% (as defined by IgG response to the following antigens) in our cohort to maximise power, which were as follows:

- BKV seropositivity for Human Polyomavirus BKV (N = 6138) – defined as positive if: antigen BK VP1 > 250.
- HHV-7 seropositivity for Human Herpesvirus-7 (N = 6106) – defined as positive if: antigen U14 > 100.
- EBV seropositivity for Epstein-Barr Virus (N = 6079) – defined as positive if: two or more of antigen VCA p18 > 250, antigen EBNA-1 > 250, antigen ZEBRA > 100, antigen EA-D > 100.

- VSV seropostivity for Varicella Zoster Virus (N = 5953) – defined as positive if: antigen gE/gl > 100.
- HHV-6 overall seropositivity for Human Herpesvirus-6 (N = 5831) – defined as positive if: one or more of antigen IE1A > 100, antigen IE1B > 100, antigen p101 k > 100.
- HSV-1 seropositivity for herpes simplex virus 1 (N = 4387) – defined as positive if: antigen 1gG > 150.
- MCV seropositivity for Merkel Cell Polyomavirus (N = 4227) – defined as positive if: antigen MC VP1 > 250.
- JCV seropositivity for Human Polyomavirus JCV (N = 3638) – defined as positive if: antigen JC VP1 > 250.
- CMV seropositivity for Human Cytomegalovirus (N = 3584) – defined as positive if: two or more of antigen pp150 Nter > 100, antigen pp 52 > 150, antigen pp 28 > 200.
- H. pylori Definition II seropositivity for *Helicobacter pylori* (N = 1845) – defined as positive if: two or more of antigen VacA > 100, antigen OMP > 170, antigen GroEL > 80, antigen Catalase > 180, antigen UreA > 130.
- T. gondii seropositivity for *Toxoplasma gondii* (N = 1694) –defined as positive if: either antigen p22 > 100 or antigen sag1 > 160.
- C. trachomatis Definition I seropositivity for *Chlamydia trachomatis* – (N = 1271) defined as positive if: antigen pGP3 > 200.
- HSV-2 seropositivity for herpes simplex virus 2 (N = 988) – defined as positive if: antigen 2mgG unique > 150.
- KSHV seropositivity for Kaposi's Sarcoma-Associated Herpesvirus (N = 471) – defined as positive if: either antigen LANA > 100 or antigen K8.1 > 175.

We tested the association between each PES and seropositive status using binomial logistic regression covaried for sex, age, age$^2$, ten SNP derived principal components, and two QC covaries related to the preparation of samples for the IgG assay. Specifically, there were two binary variables indicating a potential spill-over or samples that underwent an additional freeze-thaw cycle. In addition, amongst seropositive participants for the above pathogens we tested the association between each PES and PRS with scaled IgG response (mean fluorescence intensity) for the antigens used to define seropositivity (seroreactivity). This linear model was constructed using the same covariates as the logit above.

**Operating systems and software versions**

The unix scripts and command line inputs for this manuscript were run using either macOS Catalina version 10.15.4 or Ubuntu 18.04.5 LTS. R scripts were computed using R version 3.6.0, whilst python scripts were computed using python 3.8.3. The scripts utilised for this study will be available on GitHub: https://github.com/Williamreay/Pneumonia_meta_GWAS_drug_repurposing.

#### SUPPLEMENTARY REFERENCES


1. Reid CJ, Gould S, Harris A. Developmental expression of mucin genes in the human respiratory tract. Am J Respir Cell Mol Biol. 1997 Nov;17(5):592–8.

2. Tian C, Hromatka BS, Kiefer AK, Eriksson N, Noble SM, Tung JY, et al. Genome-wide association and HLA region fine-mapping studies identify susceptibility loci for multiple common infections. Nat Commun. 2017 19;8(1):599.

3. Durand EY, Do CB, Mountain JL, Macpherson JM. Ancestry Composition: A Novel, Efficient Pipeline for Ancestry Deconvolution [Internet]. Bioinformatics; 2014 Oct [cited 2020 Aug 31]. Available from: http://biorxiv.org/lookup/doi/10.1101/010512

4. Henn BM, Hon L, Macpherson JM, Eriksson N, Saxonov S, Pe'er I, et al. Cryptic distant relatives are common in both isolated and cosmopolitan genetic samples. PLoS ONE. 2012;7(4):e34267.

5. Zhou W, Nielsen JB, Fritsche LG, Dey R, Gabrielsen ME, Wolford BN, et al. Efficiently controlling for case-control imbalance and sample relatedness in large-scale genetic association studies. Nat Genet. 2018;50(9):1335–41.

6. Willer CJ, Li Y, Abecasis GR. METAL: fast and efficient meta-analysis of genomewide association scans. Bioinformatics. 2010 Sep 1;26(17):2190–1.

7. Watanabe K, Taskesen E, van Bochoven A, Posthuma D. Functional mapping and annotation of genetic associations with FUMA. Nat Commun. 2017 Dec;8(1):1826.

8. Piñero J, Bravo À, Queralt-Rosinach N, Gutiérrez-Sacristán A, Deu-Pons J, Centeno E, et al. DisGeNET: a comprehensive platform integrating information on human disease-associated genes and variants. Nucleic Acids Res. 2017 04;45(D1):D833–9.

9. Eppig JT. Mouse Genome Informatics (MGI) Resource: Genetic, Genomic, and Biological Knowledgebase for the Laboratory Mouse. ILAR Journal. 2017 Jul 1;58(1):17–41.

10. Bulik-Sullivan B, Finucane HK, Anttila V, Gusev A, Day FR, Loh P-R, et al. An atlas of genetic correlations across human diseases and traits. Nat Genet. 2015 Nov;47(11):1236–41.

11. Wellcome Trust Case Control Consortium, Maller JB, McVean G, Byrnes J, Vukcevic D, Palin K, et al. Bayesian refinement of association signals for 14 loci in 3 common diseases. Nat Genet. 2012 Dec;44(12):1294–301.

12. Wakefield J. Bayes factors for genome-wide association studies: comparison with P-values. Genet Epidemiol. 2009 Jan;33(1):79–86.

13. Zhu Z, Zheng Z, Zhang F, Wu Y, Trzaskowski M, Maier R, et al. Causal associations between risk factors and common diseases inferred from GWAS summary data. Nat Commun. 2018 Dec;9(1):224.

14. Liu M, Jiang Y, Wedow R, Li Y, Brazel DM, Chen F, et al. Association studies of up to 1.2 million individuals yield new insights into the genetic etiology of tobacco and alcohol use. Nat Genet. 2019;51(2):237–44.

15. de Leeuw CA, Mooij JM, Heskes T, Posthuma D. MAGMA: generalized gene-set analysis of GWAS data. PLoS Comput Biol. 2015 Apr;11(4):e1004219.

16. Liberzon A, Birger C, Thorvaldsdóttir H, Ghandi M, Mesirov JP, Tamayo P. The Molecular Signatures Database (MSigDB) hallmark gene set collection. Cell Syst. 2015 Dec 23;1(6):417–25.

17. Reay WR, Cairns MJ. Pairwise common variant meta-analyses of schizophrenia with other psychiatric disorders reveals shared and distinct gene and gene-set associations. Transl Psychiatry. 2020 Dec;10(1):134.

18. Liu Y, Xie J. Cauchy Combination Test: A Powerful Test With Analytic *p* -Value Calculation Under Arbitrary Dependency Structures. Journal of the American Statistical Association. 2020 Jan 2;115(529):393–402.

19. Liu Y, Chen S, Li Z, Morrison AC, Boerwinkle E, Lin X. ACAT: A Fast and Powerful p Value Combination Method for Rare-Variant Analysis in Sequencing Studies. Am J Hum Genet. 2019 07;104(3):410–21.

20. Talluri R, Shete S. A linkage disequilibrium-based approach to selecting disease-associated rare variants. PLoS ONE. 2013;8(7):e69226.

21. Turkmen A, Lin S. Are rare variants really independent? Genet Epidemiol. 2017;41(4):363–71.

22. Wang K, Li M, Hakonarson H. ANNOVAR: functional annotation of genetic variants from high-throughput sequencing data. Nucleic Acids Res. 2010 Sep;38(16):e164.

23. Gusev A, Ko A, Shi H, Bhatia G, Chung W, Penninx BWJH, et al. Integrative approaches for large-scale transcriptome-wide association studies. Nat Genet. 2016 Mar;48(3):245–52.

24. Giambartolomei C, Vukcevic D, Schadt EE, Franke L, Hingorani AD, Wallace C, et al. Bayesian test for colocalisation between pairs of genetic association studies using summary statistics. PLoS Genet. 2014 May;10(5):e1004383.

25. Mancuso N, Freund MK, Johnson R, Shi H, Kichaev G, Gusev A, et al. Probabilistic fine-mapping of transcriptome-wide association studies. Nat Genet. 2019;51(4):675–82.

26. Zheng J, Erzurumluoglu AM, Elsworth BL, Kemp JP, Howe L, Haycock PC, et al. LD Hub: a centralized database and web interface to perform LD score regression that maximizes the potential of summary level GWAS data for SNP heritability and genetic correlation analysis. Bioinformatics. 2017 15;33(2):272–9.

27. O'Connor LJ, Price AL. Distinguishing genetic correlation from causation across 52 diseases and complex traits. Nat Genet. 2018;50(12):1728–34.

28. Hemani G, Zheng J, Elsworth B, Wade KH, Haberland V, Baird D, et al. The MR-Base platform supports systematic causal inference across the human phenome. eLife. 2018 May 30;7:e34408.

29. Willer CJ, Schmidt EM, Sengupta S, Peloso GM, Gustafsson S, Kanoni S, et al. Discovery and refinement of loci associated with lipid levels. Nat Genet. 2013 Nov;45(11):1274–83.

30. Freshour S, Kiwala S, Cotto KC, Coffman AC, McMichael JF, Song J, et al. Integration of the Drug-Gene Interaction Database (DGIdb) with open crowdsource efforts [Internet]. Bioinformatics; 2020 Sep [cited 2020 Nov 4]. Available from: http://biorxiv.org/lookup/doi/10.1101/2020.09.18.301721

31. Burgess S, Butterworth A, Thompson SG. Mendelian randomization analysis with multiple genetic variants using summarized data. Genet Epidemiol. 2013 Nov;37(7):658–65.

32. Bowden J, Hemani G, Davey Smith G. Invited Commentary: Detecting Individual and Global Horizontal Pleiotropy in Mendelian Randomization-A Job for the Humble Heterogeneity Statistic? Am J Epidemiol. 2018 01;187(12):2681–5.

33. Bowden J, Davey Smith G, Haycock PC, Burgess S. Consistent Estimation in Mendelian Randomization with Some Invalid Instruments Using a Weighted Median Estimator. Genet Epidemiol. 2016 May;40(4):304–14.

34. Bowden J, Davey Smith G, Burgess S. Mendelian randomization with invalid instruments: effect estimation and bias detection through Egger regression. Int J Epidemiol. 2015 Apr;44(2):512–25.

35. Hartwig FP, Davey Smith G, Bowden J. Robust inference in summary data Mendelian randomization via the zero modal pleiotropy assumption. Int J Epidemiol. 2017 01;46(6):1985–98.

36. Verbanck M, Chen C-Y, Neale B, Do R. Detection of widespread horizontal pleiotropy in causal relationships inferred from Mendelian randomization between complex traits and diseases. Nat Genet. 2018;50(5):693–8.

37. Reay WR, Atkins JR, Carr VJ, Green MJ, Cairns MJ. Pharmacological enrichment of polygenic risk for precision medicine in complex disorders. Sci Rep. 2020 Jan 21;10(1):879.

38. Reay WR, Shair SE, Geaghan MP, Riveros C, Holiday EG, McEvoy MA, et al. Genetically informed precision drug repurposing for lung function and implications for respiratory infection [Internet]. Respiratory Medicine; 2020 Jun [cited 2020 Sep 8]. Available from: http://medrxiv.org/lookup/doi/10.1101/2020.06.25.20139816

39. Liao Y, Wang J, Jaehnig EJ, Shi Z, Zhang B. WebGestalt 2019: gene set analysis toolkit with revamped UIs and APIs. Nucleic Acids Res. 2019 02;47(W1):W199–205.

40. Sudlow C, Gallacher J, Allen N, Beral V, Burton P, Danesh J, et al. UK biobank: an open access resource for identifying the causes of a wide range of complex diseases of middle and old age. PLoS Med. 2015 Mar;12(3):e1001779.

41. Bycroft C, Freeman C, Petkova D, Band G, Elliott LT, Sharp K, et al. The UK Biobank resource with deep phenotyping and genomic data. Nature. 2018 Oct;562(7726):203–9.

42. Abraham G, Qiu Y, Inouye M. FlashPCA2: principal component analysis of Biobank-scale genotype datasets. Stegle O, editor. Bioinformatics. 2017 Sep 1;33(17):2776–8.

43. Price AL, Weale ME, Patterson N, Myers SR, Need AC, Shianna KV, et al. Long-range LD can confound genome scans in admixed populations. Am J Hum Genet. 2008 Jul;83(1):132–5; author reply 135-139.

44. Mentzer AJ, Brenner N, Allen N, Littlejohns TJ, Chong AY, Cortes A, et al. Identification of host-pathogen-disease relationships using a scalable Multiplex Serology platform in UK Biobank [Internet]. Infectious Diseases (except HIV/AIDS); 2019 Aug [cited 2020 Dec 3]. Available from: http://medrxiv.org/lookup/doi/10.1101/19004960
